# A statistical calibration framework for subnational health surveillance from routine data and survey audits

**DOI:** 10.64898/2026.10.27.26366475

**Authors:** Adrien Allorant, Li-Chun Zhang

## Abstract

In many low- and middle-income countries, where civil registration and vital statistics remain incomplete, decision makers rely on estimates derived from large international household surveys. Those surveys are rarely powered for frequent subnational estimates, and the contraction in international aid has made their future uncertain. Small area estimation has supplemented them with increasingly available routine health records and satellite imagery to improve their precision, but the survey estimate remains the response variable. We propose a statistical calibration framework that inverts the roles of the two data sources, whereby the routine records provide a biased baseline estimator continuously over time at the subnational level of interest, while a probability survey is used intermittently to audit and adjust for the selection bias of the direct register estimator. We develop the corresponding modelling and estimation methods, including diagnostic and analysis tools for the transport assumption that the out-of-survey subpopulations and time points require. The proposed framework is applied in four African countries to estimate undiagnosed HIV positivity by district and the selection bias by age and sex, and we explain how it applies to other health surveillance needs with similar data settings.

## 1 Introduction

Subnational estimates of health service coverage reveal where a health system is failing to reach people and guide the resources that would close the gap (Tanahashi, 1978; Faye et al., 2020). Most low- and lower-middle-income countries (LMICs) have relied on periodic household surveys to measure their health systems’ reach and the gaps within it (AbouZahr et al., 2015). Over the past two decades, sustained investment has built a second source, the routine health information system (RHIS), which records every interaction between a patient and the health system at the point of service provision (Maïga et al., 2019).

International large-scale household surveys conducted in LMICs, such as the Demographic and Health Surveys (DHS), or the Multiple Indicator Cluster Surveys, use a stratified two-stage cluster sampling design to provide nationally representative estimates of various health outcomes (ICF, 2025; UNICEF, 2025). Because every household has a known probability of selection, the estimates are representative of the whole population by construction and their uncertainty is quantified from the design alone (Neyman, 1934). Yet their sample sizes are usually meant for reliable national and provincial estimates. They are therefore too sparse to resolve the district and age–sex detail used in programme decisions (Mercer et al., 2015). These surveys are also costly, time-consuming and logistically challenging. They have therefore been financed almost entirely by international donors and are conducted in a given country only about every five years, with the DHS programme as a whole funded at roughly US$47 million a year by a single donor (Wahl et al., 2025). This dependence on external funding crystallised in 2025, when the DHS programme lost its principal funder with the dissolution of USAID and has since continued only in part, on interim philanthropic funding, while the Population-based HIV Impact Assessment (PHIA) surveys depend on PEPFAR, whose own funding became uncertain in the same year (The White House, 2025; KFF, 2025; CSIS, 2025).

RHIS record patients’ contact with the health system. Every test performed, every vaccine administered and every antenatal visit is logged by period and facility, so the accumulated records amount to a continuously refreshed register of the population that comes into contact with services (Lippeveld, Sauerborn and Bodart, 2000; AbouZahr and Boerma, 2005). The resulting dataset is granular, rich and timely, resolved by facility and month, the spatial and temporal resolution of resource allocation and action in health systems (Hotchkiss, Diana and Foreit, 2012; Ndabarora, Chipps and Uys, 2014; Wickremasinghe et al., 2016; Hoxha et al., 2022). Long-standing concerns about the quality of routine records (Mbondji et al., 2014) have been tempered on two fronts. Interventions have strengthened district-level reporting (Lee et al., 2021; Muhoza et al., 2022; Aron et al., 2024), and national systems have consolidated on shared open platforms, with DHIS2 adopted as a locally owned health information system in more than 80 countries (HISP Centre, 2026). Assessments of reporting completeness and internal consistency, once ad hoc, are now conducted routinely on national systems (Aron et al., 2024; Dhodho et al., 2026).

Collecting this information is not itself cheap, absorbing up to a third of health workers’ time in African countries (Kruk et al., 2018). But, using these records to draw estimates of health services coverage or for surveillance costs the statistical system almost nothing, because the records already exist as a by-product of care and belong to the Ministry of Health that collects them (Wagenaar et al., 2016). However, routine records capture only a selected sample of the population; the people who come into contact with services. Framed statistically, the register is a very large non-probability sample, because people who present for a service are not a random sample of those who need it and the propensity to seek care is correlated with the outcome being measured. Meng (2018) highlighted that the error of an estimate from such a sample is governed by the correlation between inclusion and the outcome, scaled by the size of the population. For a fixed inclusion fraction that error does not shrink as the population grows, while the nominal standard error does, so a very large register can be precise and wrong at once. Vaccine uptake in the United States was overestimated from unrepresentative big surveys through exactly that mechanism (Bradley et al., 2021).

Survey data and routine records are two complementary sources, with strengths that fall in different places. Combining them is a well-known idea in small-area estimation (SAE). Fay and Herriot (1979) estimated per-capita income for small places in the United States, combining direct estimates from the census sample with regression predictions built on administrative tax records and housing data, each weighted by its precision. Direct survey estimates for fine domains, such as administrative subdivisions (“district”) or age and sex disaggregation, are too noisy to use on their own. The area-level approach therefore treats each direct estimate as a noisy measurement of the true area parameter, with sampling variance known from the design. It then links the true quantities across areas through auxiliary data in a regression model (Spjøtvoll and Thomsen, 1987; Pfeffermann, 2013; Rao and Molina, 2015). The empirical best predictor then pulls each area’s direct estimate toward its regression prediction, with a weight set by the relative precision of the two, and such estimators have become common in the production of official statistics (Tzavidis et al., 2018). The framework has been extended to auxiliary sources that are themselves error-prone, with the noise in the covariates accounted for in measurement-error models (Ybarra and Lohr, 2008). Small-area methods now underpin subnational estimation across public health, from all-cause mortality and life expectancy (GBD US Health Disparities Collaborators, 2022) to child mortality (Dwyer-Lindgren et al., 2013; Li et al., 2019; Godwin and Wakefield, 2021), vaccination coverage (Dong and Wakefield, 2021), tuberculosis (Ross et al., 2018; Allorant et al., 2022), and health service readiness (Allorant et al., 2023). The Naomi model (Eaton et al., 2021; Esra et al., 2024), used by more than forty African countries for their official subnational HIV statistics, also takes the survey as the response and the routine data as auxiliary covariates with which survey estimates are extended to unsampled areas.

However, SAE as such is limited by the availability of survey data, and it is impossible in the years without a survey. In this article, we invert the roles of survey and routine data, such that the register provides a baseline estimator that is timely, granular and state-owned, and the survey becomes a periodic audit whose task is to estimate the register’s selection bias to account for it. National statistical offices already draw official estimates from non-randomised sources, with the selection mechanism modelled and corrected (Zhang, 2012, 2015). Both Norway and Sweden implemented a register-based census in 2011, checked against an independent quality survey (Hendriks, 2021; Axelson et al., 2021). Statistics Netherlands compiles a “virtual census” by integrating registers with its labour force survey (Schulte Nordholt, 2018; Bakker, van Rooijen and van Toor, 2014). Ireland’s Central Statistics Office produces annual population estimates from administrative data alone (Dunne and Zhang, 2024). Two features set our problem apart. First, inclusion in the register is informative, driven by care-seeking, which is correlated with the condition being measured. A selection effect must therefore be estimated from the audit (Zhang, 2019). Second, the domain is subnational disease surveillance in LMICs, unlike the register-based statistics that exist in high-income countries.

We develop the calibration framework in general form and apply it to the estimation of undiagnosed HIV positivity, with selection resolved by age and sex and prediction at district level. The four countries are Mozambique, Zimbabwe, Malawi and Lesotho, all with a high HIV burden in eastern and southern Africa. The application is consequential: reaching people living with HIV who are not yet aware of their status, the first of the 95–95–95 targets set by the Joint United Nations Programme on HIV/AIDS (UNAIDS), is a crucial entry point to treatment (Maheu-Giroux et al., 2019b), and a prerequisite for ending the AIDS epidemic by 2030 (United Nations, 2015). It is also a natural illustration for our proposed framework, since the two required ingredients are in place, a mature routine health information system that records testing and treatment, and recent household surveys that overlap in space and time. The approach has two principal advantages, (a) timeliness, since estimates are available at the cadence of the register, and (b) sovereignty, as estimates are derived from routine systems run and owned by ministries of health in the respective countries.

The paper is organised as follows. Section 2 introduces the setting, data and estimand. Section 3 develops the calibration model in general form and applies it to HIV surveillance. Section 4 develops the estimation method, the associated uncertainty evaluation and a model diagnostic. Section 5 applies the framework to the four countries. In Section 6, a known-truth simulation fixes the conditions under which the calibrated framework improves on current practice, and provides the analysis tools for future applications in similar settings. We discuss the results and conclude in Section 7. Appendices A to C are in the Web-based supporting materials.

## 2 Setting, data and estimand

### 2.1 Two complementary sources

The first data source is routine HIV testing, provided by the national ministries of health from their routine health information systems (e.g. DHIS2) as facility-level testing counts. We aggregate these to the district *d* and age–sex stratum *g*. We focus on two variables: the number of HIV tests performed,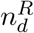, and the number returning a positive result, 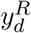, whose ratio is the HIV testing positivity 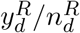. For each country we use the single year of routine data that matches the survey year. National annual test volumes range from about 0.3 to 3.7 million (Table 3). Relative to its own population, the people who present for a test, the register is near complete. The target population of the estimand, defined in Section 2.2, is everyone not yet diagnosed, and the register includes a selected subset of it.

The second source is the Population-based HIV Impact Assessment (PHIA), a household survey fielded in each country between 2020 and 2021: INSIDA 2021 in Mozambique, LePHIA 2020 in Lesotho, ZIMPHIA 2020 in Zimbabwe and MPHIA 2020–21 in Malawi (Justman et al., 2018). Each employs a stratified two-stage probability sample, of enumeration areas and then of households, covering adults aged 15 and over. Consenting participants give a blood sample, from which HIV status is determined by validated biomarker and antiretroviral metabolites are assayed. They also report their prior testing and diagnosis, a measure of reported HIV status and awareness of HIV status. Biomarker-tested sample sizes range from about 14,500 to 22,700 adults and household response exceeded 88% in every country (Table 3). The cascade summary of Table 1 is restricted to ages 15–64, following the PHIA country reports. PHIA is designed to be precise at national and provincial (admin-1) level; direct estimates at the district (admin-2) may be too unreliable or nonexistent in many areas.

**Table 1.** The first 95: awareness of HIV-positive status among people living with HIV, by country and sex. Per cent (95% CI).

| Country | National | Male | Female |
| --- | --- | --- | --- |
| Lesotho | 89.9 (88.7–91.0) | 87.7 (85.2–89.8) | 91.2 (90.0–92.3) |
| Malawi | 88.4 (86.7–89.9) | 85.2 (82.1–87.8) | 90.4 (88.5–92.0) |
| Mozambique | 71.3 (68.0–74.4) | 67.5 (61.6–72.8) | 73.4 (70.0–76.6) |
| Zimbabwe | 86.6 (84.9–88.2) | 83.7 (80.4–86.5) | 88.4 (86.8–89.8) |

Source: PHIA microdata; awareness = self-report or detectable antiretrovirals; design-adjusted (strata, PSU, biomarker sampling weight); adults aged 15–64 living with HIV. National second-95 (ART given aware) and third-95 (viral suppression given ART), for context: Lesotho 96.8 / 91.1; Malawi 97.8 / 96.9; Mozambique 96.3 / 89.3; Zimbabwe 96.8 / 90.1.

### 2.2 The HIV care cascade and the choice of estimand

The HIV care cascade is the sequence of stages from acquisition through awareness, treatment (antiretroviral therapy, ART) and viral suppression, formalised by the UNAIDS 95–95–95 targets. We illustrate the framework on the first 95, knowledge of status, which across these four settings is the stage where achievement is lowest and so the gap that most constrains the public health response (Table 1). Our proposed approach could apply to the second 95 (treatment) and potentially the third (viral suppression), if recorded in both the survey and the routine system. Across the four countries, awareness is lowest in Mozambique, and lower in men than women throughout, consistent with disparities in HIV testing and treatment documented across African countries (Allorant et al., 2025).

The raw positivity *y*^*R*^*/n*^*R*^ is a different quantity from prevalence. Prevalence is a share of the whole population, whereas (*y*^*R*^, *n*^*R*^) here refer only to those who presented for a test, a group that is a selected subset of the population. Total prevalence decomposes as

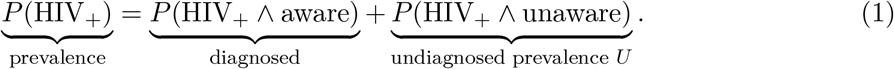

Those presenting for a routine HIV test are overwhelmingly people who do not yet know their status, because under current WHO guidance and national testing algorithms people already diagnosed should not re-test for diagnosis (WHO, 2021). Empirically, policy and practice can diverge, and people living with HIV and aware of their status may occasionally re-test through antenatal re-testing during a pregnancy, non-disclosure of status or unlinked records. The RHIS positivity therefore targets people not yet diagnosed, as a fraction testing positive among those tested, which differs from the population prevalence. We take as the estimand the undiagnosed positivity, which is the HIV positivity among people not yet diagnosed in district *d* and stratum *g*, denoted by

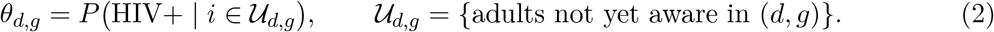

Equivalently, in a population of size *N* with *N*_*y*_ living with HIV of whom *K* are aware,

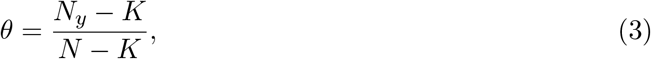

the counterfactual positivity that would be observed if every adult not yet diagnosed were tested. Re-testing by people already diagnosed is excluded from the estimand by construction, though not from the register, and its effect on the offset is bounded in Section 7.2. Writing *a* = *K/N*_*y*_ for awareness of status, the first 95 in Table 1, and *q* = *N*_*y*_*/N* for prevalence, we have

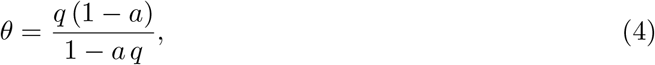

which falls as awareness rises at fixed prevalence. Table 2 reports prevalence, awareness, undiagnosed prevalence and 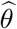 for the four surveys, design-weighted over ages 15–64. The estimate 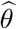 ranges from 1.1% in Malawi to 4.0% in Mozambique. Mozambique has a lower prevalence than Lesotho and the highest undiagnosed positivity of the four, because its awareness is the lowest.

**Table 2.**
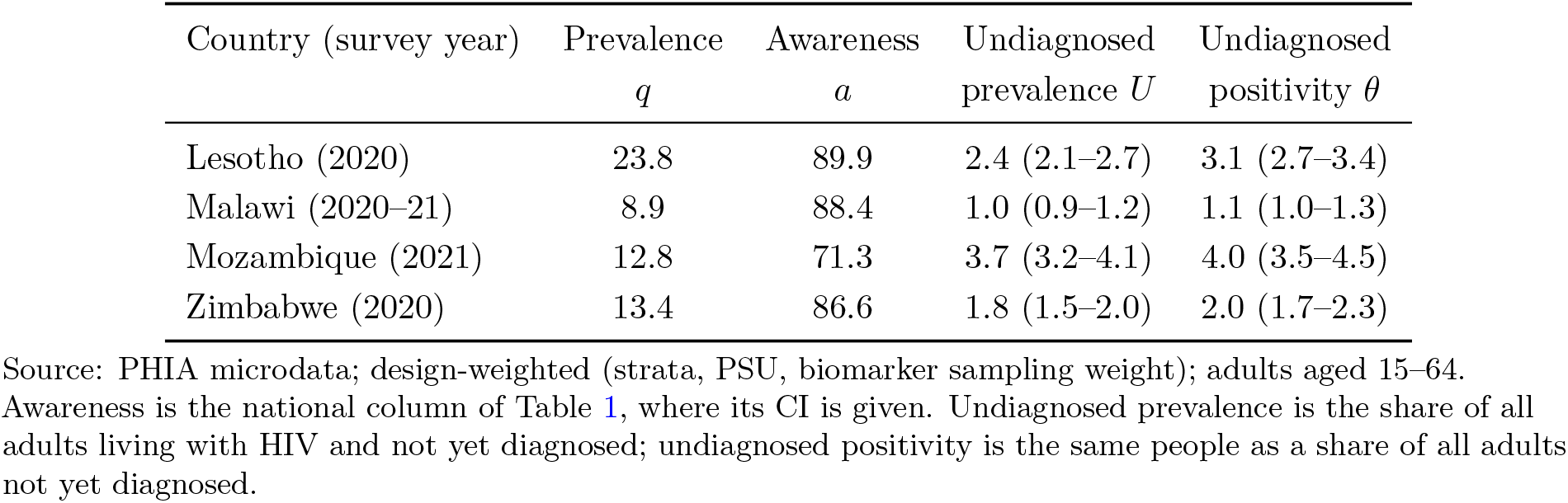
Prevalence *q*, awareness of status *a*, undiagnosed prevalence *U* = *q*(1−*a*) and undiagnosed positivity *θ* = *U/*(1 − *aq*), by country and survey year. Per cent (95% CI).

| Country (survey year) | Prevalence<br>$q$ | Awareness<br>$a$ | Undiagnosed<br>prevalence $U$ | Undiagnosed<br>positivity $\theta$ |
| --- | --- | --- | --- | --- |
| Lesotho (2020) | 23.8 | 89.9 | 2.4 (2.1–2.7) | 3.1 (2.7–3.4) |
| Malawi (2020–21) | 8.9 | 88.4 | 1.0 (0.9–1.2) | 1.1 (1.0–1.3) |
| Mozambique (2021) | 12.8 | 71.3 | 3.7 (3.2–4.1) | 4.0 (3.5–4.5) |
| Zimbabwe (2020) | 13.4 | 86.6 | 1.8 (1.5–2.0) | 2.0 (1.7–2.3) |

Source: PHIA microdata; design-weighted (strata, PSU, biomarker sampling weight); adults aged 15–64. Awareness is the national column of Table 1, where its CI is given. Undiagnosed prevalence is the share of all adults living with HIV and not yet diagnosed; undiagnosed positivity is the same people as a share of all adults not yet diagnosed.

Taking undiagnosed positivity *θ* as the estimand matches the quantity identifiable from the register. A population prevalence would additionally require the size of the diagnosed population and of the district’s adult population, because the already-diagnosed do not re-enter routine testing and the register records neither. The quantity *θ* is also the planning quantity of a testing programme, the expected yield among the people reached by the programme, and the bias of the observed register positivity is additive on the log-odds scale, as we show in Section 3.

The target *θ* is directly measurable in the PHIA survey. Each participant provides a blood sample for biomarker-confirmed HIV status and answers questions on prior testing and diagnosis, and counts as aware on a self-reported prior diagnosis or on detectable antiretroviral metabolites. Those who know their status but do not report it are identified from the metabolites. Setting aside those already aware of their status, *θ* is the design-weighted proportion with HIV among people not yet diagnosed.

We shall apply the framework to these four countries, each with a recent PHIA survey paired to a matching-year RHIS extract (Table 3).

**Table 3.**
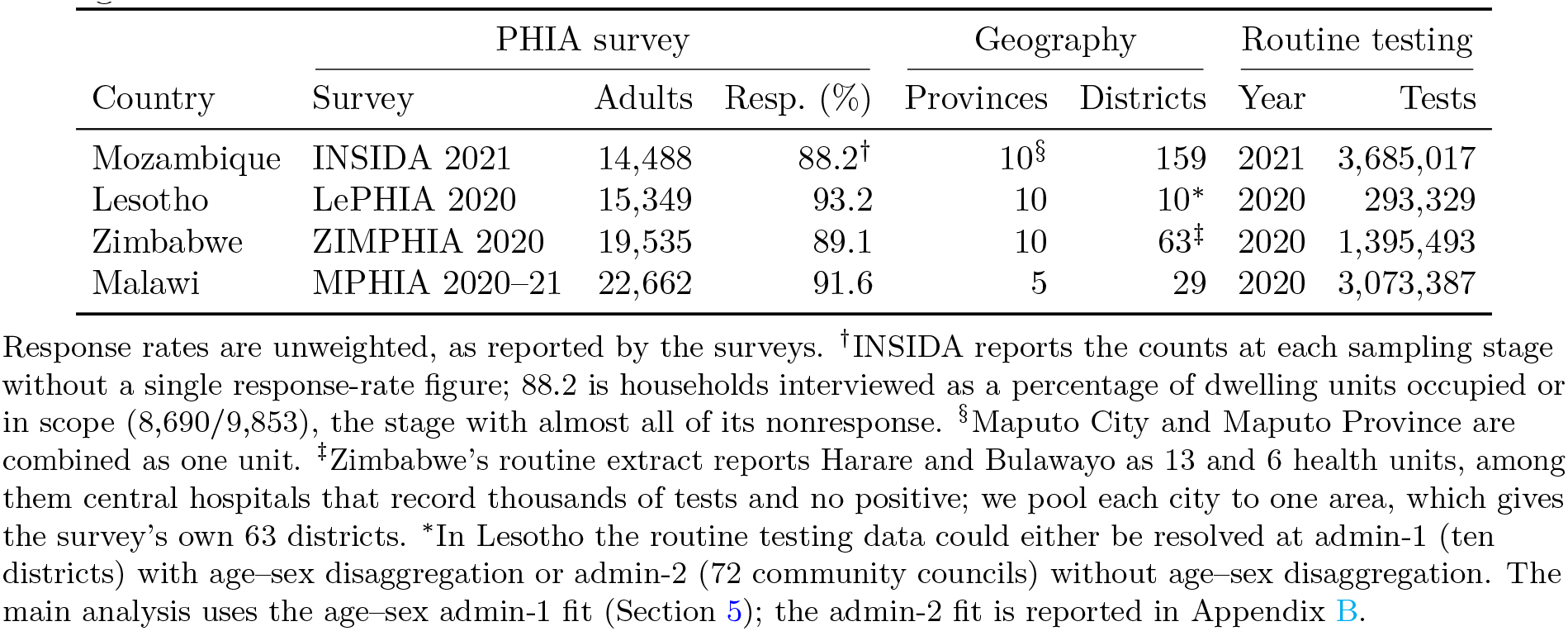
Data sources. Adults are the biomarker-tested survey sample and response is the household response rate. Provinces are the survey’s design domains: admin-1 provinces in Mozambique and Zimbabwe, the five health zones in Malawi, and in Lesotho its ten districts, which are its admin-1 units. Source: PHIA survey reports and ministry-of-health routine (RHIS) testing data.

## 3 The statistical calibration framework

### 3.1 A general formulation

The formulation applies wherever a biased estimator is available from a register and an unbiased one from a probability survey for the same subnational target. For a population cell *i*, let *θ*_*i*_ be the target parameter, *z*_*i*_ a register statistic and *y*_*i*_ a survey statistic, with

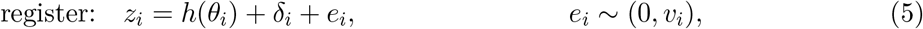

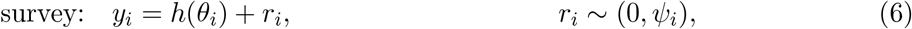

where *h* is a transformation chosen to make the register’s bias additive, *v*_*i*_ is the variance of *z*_*i*_, *ψ*_*i*_ that of *y*_*i*_, and *δ*_*i*_ is the bias of *z*_*i*_ for *h*(*θ*_*i*_). Given any estimator 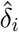, the target follows from the register as

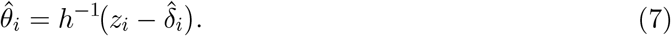

Where cell *i* is sampled by the survey, an unbiased direct estimator of *δ* is 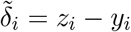, since 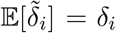. Using 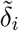 in (7) returns the survey estimate itself, 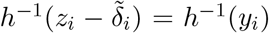. We use statistical modelling to obtain a more precise estimate.

Small-area models are one route to it. A Fay–Herriot model (Fay and Herriot, 1979) of *y*_*i*_ in terms of (6) conditions on *z*_*i*_ as a fixed covariate,

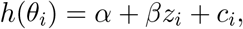

with random effects *c*_*i*_ for the discrepancy between *h*(*θ*_*i*_) and the fixed-effects predictor *α* + *βz*_*i*_. Equation (5) gives *h*(*θ*_*i*_) = E[*z*_*i*_] − *δ*_*i*_, so register and target are connected directly. Cancelling *h*(*θ*_*i*_) between (5) and (6) leaves

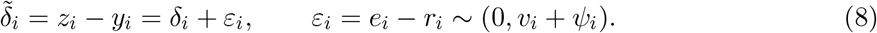

The register error *e*_*i*_ and the survey sampling error *r*_*i*_ are independent of each other.

Covariate structure makes (8) worth fitting. Without it, a weighted least squares estimator of the vector *δ* returns the direct estimate 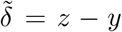 under any diagonal weight matrix *W*, since 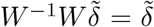. Spatial structure can be added where it is needed in an application, and temporal effects where (*z*_*i*_, *y*_*i*_) are observed at several time points. Random-effects modelling of *z*_*i*_ = *y*_*i*_ + *δ*_*i*_ + *ε*_*i*_, or of *y*_*i*_ = *z*_*i*_ − *δ*_*i*_ − *ε*_*i*_, can be read as a measurement-error Fay–Herriot model (Ybarra and Lohr, 2008) with *y*_*i*_, or *z*_*i*_, as the random covariate. The corresponding regression coefficient is fixed at 1, so the model estimates nothing beyond the difference (8), and we work with the difference.

In the formulation (8) we move the modelling from the target *θ*_*i*_ to the register bias *δ*_*i*_, and the statistical calibration perspective of this paper is defined by that move. Once 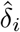 is estimated at a time when (*z*_*i*_, *y*_*i*_) are observed together, (7) can be updated for later periods with a refreshed register alone. Undiagnosed HIV positivity is an instance of that setting.

The scale *h* is the logit for a proportion, logit(*p*) = log{*p/*(1 − *p*)}, such as the undiagnosed HIV positivity developed below, and the log for a rate or a denominator-limited count, such as tuberculosis case notification or childhood immunisation coverage.

### 3.2 Application to HIV: the log-odds identity

For HIV surveillance let 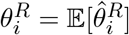 be the test positivity, where 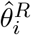 is the proportion of positive HIV tests among all tests recorded in the RHIS for cell *i*. This is related to, and different from, the undiagnosed positivity *θ*_*i*_ defined as the target in Section 2.2.

Among the people in cell *i* not yet aware of their status, let *π*_1*i*_ be the probability of taking a test given the person has HIV, and *π*_0*i*_ that probability given the person does not have HIV. The odds of a positive test in the register follow as

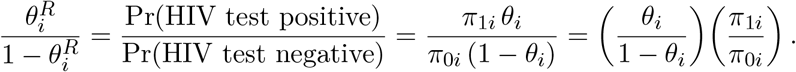

Writing *h*(*θ*) = logit(*θ*) = log *θ* − log(1 − *θ*) for the log-odds gives the calibration identity

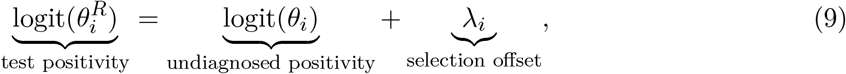

where *λ*_*i*_ is the register-selection offset, the logarithm of the relative test propensity (RTP),

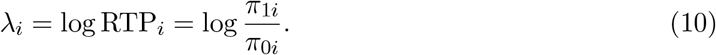

Identity (9) holds for any register proportion where the register covers a subset of the target population and the probability of entering it differs by the binary status of interest. In HIV surveillance the target population is everyone not yet diagnosed, some of whom are recorded in the register at an HIV test. The bias of 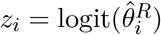 is additive on the log-odds scale, that is, the general bias *δ*_*i*_ of (5) reduces to the additive selection offset *λ*_*i*_ of (10). Statistical calibration here requires an estimate of *λ*_*i*_. The register counts tests rather than people. Reading *π*_1*i*_ and *π*_0*i*_ as the expected number of recorded tests per person over the period, among people not yet diagnosed with and without HIV, leaves the derivation unchanged, since the odds of a positive record are then the ratio of expected positive to expected negative tests. Repeat testing among people without HIV, such as antenatal re-testing late in pregnancy or periodic testing in key populations, raises *π*_0*i*_, lowers *λ*_*i*_ and is part of the offset.

### 3.3 Covariate structure and the transport assumption

The cells of interest are a district crossed with a sex and age group, *i* = (*d, g*), with age groups 15–24, 25–49 and 50 and over. Table 3 gives the number of districts in each country, from 10 to 159. The survey estimate *y*_*i*_ is not available in every cell, so structure has to be placed on the offset in the cells sampled by the survey to determine the offset in the remaining cells. In SAE terminology, the regression function of (8) needs a synthetic part.

Three structures are available at this indexing. Under a single offset per group, 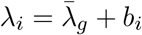 with 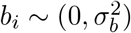, and the group mean 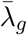 is transported to every cell without an estimate of *b*_*i*_. Under main effects, the offset is the sum of a district term and a group term, so the age–sex pattern of selection is the same in every district and the district pattern is the same in every group. Under broad areas, each cell of interest is nested in a province by group cell with survey support, and we write *p*(*d*) for the province containing district *d*,

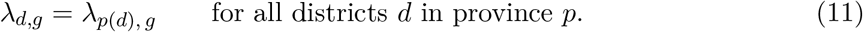

Broad-area structures of this kind, with the cells of interest nested in estimable broad-area cells, are common in small-area estimation.

We decide between the second and the third options by reading parameter counts against the survey’s sample support. Main effects need 164 parameters in Mozambique and 15 in Lesotho, whose surveys are of about the same size (Table 3), 68 in Zimbabwe and 30 in Malawi. Broad areas need 60 in each of the three countries with six groups and 10 in Malawi, whose register records sex without age. Not every district is present in the survey or has adequate sample support in it. The choice follows from the case of Lesotho. Its districts and provinces coincide, so the 60-parameter broad-area structure includes the province by group interaction omitted from the 15-parameter main-effects structure, and the latter involves no transport beyond the main effects. We adopt (11) throughout.

Efficiency beyond direct estimation of each *λ*_*p,g*_ needs a covariate in addition to (*p, g*), for the reason given in Section 3.1. Writing *η* = logit *θ* for the working scale,

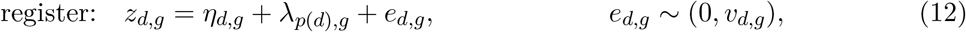

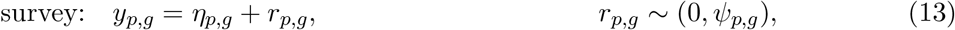

where 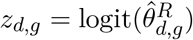 comes from the observed proportion of positive tests in the register, and *y*_*p,g*_ targets logit(*θ*_*p,g*_) of the broad-area cell. The variance of *z*_*d,g*_ follows by linearising 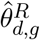 with respect to the binomial distribution,

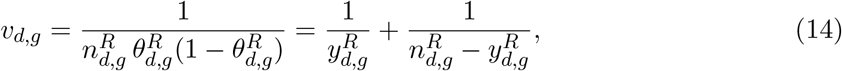

with the second equality exact at 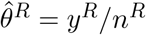, where it reduces to Woolf’s variance for a log-odds (Woolf, 1955). It is small because routine-data denominators are large. The variance of *y*_*p,g*_ follows by linearising 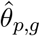, whose variance is calculated with respect to the sampling design. Both are treated as known, as is usual for Fay–Herriot models.

The survey estimator of (6) is therefore specified at province level, matching the target offsets under (11) and better supported by the sample than a district-level *y*_*d,g*_. The register is specified at district level, which alone makes a district-level prediction available.

Under (11), differencing (12) and (13) turns the general formulation (8) into a Fay–Herriot model,

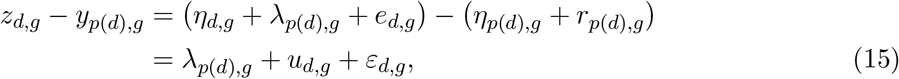

Where

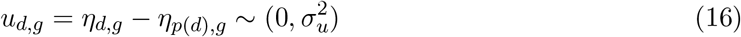

is a random effect for the discrepancy between the district target and its broad-area value, except in Lesotho where (*d, g*) and (*p, g*) coincide and *u*_*d,g*_ ≡ 0, and

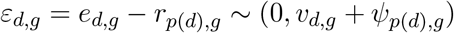

combines the register and survey sampling errors. The register error and the survey error are independent of each other, but *ε*_*d,g*_ and *ε*_*l,g*_ nonetheless share the same *r*_*p*(*d*),*g*_ whenever districts *d* and *l* belong to one province. Let *D*_*p,g*_ = *{*(*d, g*) : ∃ *y*_*d,g*_, *p*(*d*) = *p}* collect the sample districts of province *p*. For *d* ≠ *l* ∈ *D*_*p,g*_,

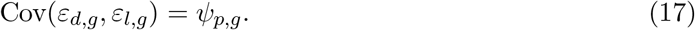

Model (15) assumes independent errors and random effects at (*d, g*) and (*p, g*), apart from the within-province covariance (17). In Appendix B we examine whether spatial correlation is worth adding and find no strong evidence for it. The application uses the covariate structure of (15) alone.

## 4 Estimation

In model (15) each offset *λ*_*p,g*_ is a fixed effect and each *u*_*d,g*_ a random effect with unknown variance 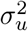. We estimate them as follows, and then devise a diagnostic for the transport assumption (11) that underlies both.

### 4.1 The selection offsets

The best linear unbiased estimator of *λ*_*p,g*_ is the generalised least squares (GLS) estimator given the variance components. Let 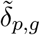 be the vector of *z*_*d,g*_ − *y*_*p*(*d*),*g*_ over *d* ∈ *D*_*p,g*_. Its variance-covariance matrix is

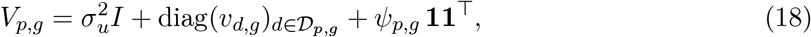

with *I* the identity matrix and **1** the unit vector, both of dimension *n*_*p,g*_ = |*D*_*p,g*_|, the off-diagonal entries arising from (17). The estimator follows as

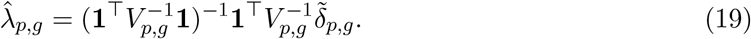

By the derivation in Appendix A.1, the associated variance simplifies to

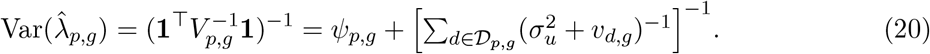

Only the bracketed term falls as more districts are added. Every district in the cell is differenced against the same survey estimate, so all of them share one survey error and *ψ*_*p,g*_ enters undivided by *n*_*p,g*_. Replacing 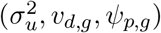 by their estimates gives the empirical GLS estimate of *λ*_*p,g*_ together with its plug-in variance.

The calibrated register estimator of *θ*_*d,g*_, in the general form (7), is

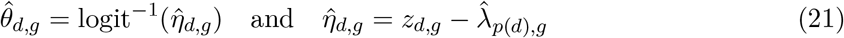

under the transport assumption (11). The prediction variance of 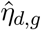 is approximately

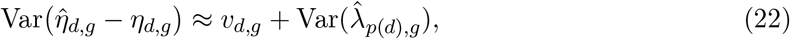

the register variance plus the survey calibration variance. Where *v*_*d,g*_ is small the second term dominates. Equation (22) omits the term 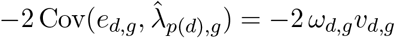, with *ω*_*d,g*_ the weight of district *d* in (19). That covariance is positive and the omitted term is negative, so the reported variance exceeds the exact prediction variance by 2 *ω*_*d,g*_*v*_*d,g*_ (Appendix A.1).

Equation (22) is the prediction variance under the transport assumption (11). Where the offset of district *d* departs from its province value by *τ*_*d,g*_ of variance 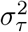, that variance adds to it, and the district intervals we report include the estimate 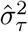 of Section 4.3 (Table S5). We report 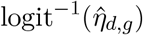 with the interval built on the log-odds scale and back-transformed. Because logit^−1^ is non-linear, the point estimate is the median of *θ*_*d,g*_ under a symmetric error on the log-odds scale, and it is within 0.1 percentage points of the mean at the median district in every country.

In a year *t* without a survey, *h* = *t − t*_0_ years after the last one, the calibrated estimate is 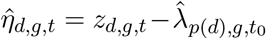, and its error against 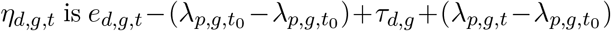: the register noise of year *t*, the calibration error of the survey round, the failure of the offset to transport, and the movement of the offset since the survey. Writing that movement as a common drift of *µ* per year plus area-level diffusion of variance 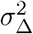 per year,

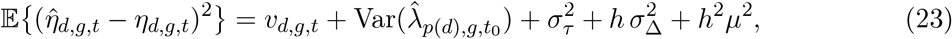

and a 95% interval for *θ* is 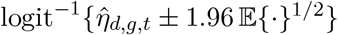. The register term is that of year *t*, computed from that year’s counts by (14), and is independent of 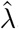, so the covariance discussed above does not arise. The common drift enters as a squared bias, since a programme holding 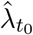 fixed cannot remove it, and (23) is therefore a mean squared error. The drift components are estimated from the two survey rounds of Section 5.2, annualised separately for the level shift and the area diffusion: on the pooled three-country data 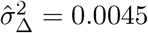 and 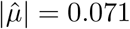 per year on the log-odds scale. Table S6 in Appendix B gives the resulting intervals.

Had one ignored the covariances (17) and applied weighted least squares (WLS) with the variance-covariance matrix Diag(*V*_*p,g*_), the WLS estimator would have remained unbiased under (11), but the associated variance would have been underestimated at 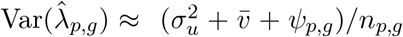, with 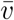 the mean of *v*_*d,g*_ over *D*_*p,g*_. Only the bracketed term of (20) is scaled by *n*_*p,g*_. Assuming independent 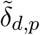 and applying the WLS variance estimator would have narrowed the standard error of the national selection offset by a factor of 1.39 to 1.74 across the four countries at the admin-2 fit (Appendix A.4), so the WLS should be avoided.

GLS is standard in small-area applications, and parametric methods that assume normally distributed errors are common alongside it. We compare GLS with two such methods in Appendix A.4, after describing them in Appendices A.2 and A.3. The first is Template Model Builder (TMB) by Kristensen et al. (2016), which estimates 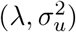 from a marginal likelihood. The second is the integrated nested Laplace approximation (INLA) of Rue et al. (2009), a fully Bayesian technique for latent Gaussian models. The estimates of *λ*_*p,g*_ are nearly equal across all three. The application reports the GLS results.

### 4.2 The random effects

Under the Fay–Herriot model (15), the best linear unbiased predictor of the vector *u*_*p,g*_ = (*u*_*d,g*_)*d*∈*D*_*p,g*_, given the variance components, is

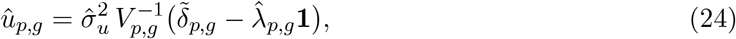

with 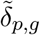 as in Section 4.1. The covariance between *u*_*p,g*_ and 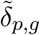 is 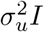, since the shared survey error belongs to the residual and not to the random effect. Replacing the variance components by their estimates gives the empirical best linear unbiased predictor (EBLUP) of *u*_*d,g*_. A predictor of *η*_*d,g*_ for *d* ∈ *D*_*p,g*_ follows from (16) as

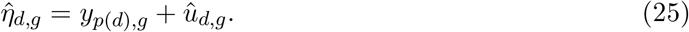

Treating *y*_*p*(*d*),*g*_ as a fixed covariate and (15) as a standard Fay–Herriot model turns (25) into a convex combination of the calibrated register (21) and the survey estimate for the broad-area cell, such that 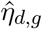 by (25) can be rewritten as

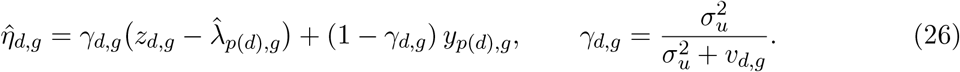

The generalised least squares estimator (19) makes the residual orthogonal to the unit vector in the metric of *V*_*p,g*_, so 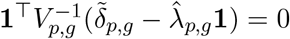 and the shared survey term drops out of (24), leaving the elementwise form above. The shrinkage factor *γ*_*d,g*_ is close to one where a register cell count 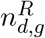 is large and falls where it is thin.

The predictor 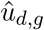 is not needed for the calibrated register estimator (21). Although (25) uses it explicitly, it depends on the same transport assumption through the definition of *u*_*d,g*_ in (16) and (15). In other words, allowing *λ*_*d,g*_ and *λ*_*p*(*d*),*g*_ to differ, we have

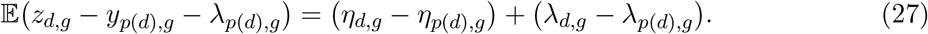

Writing *w*_*d,g*_ for the first term and *τ*_*d,g*_ for the second, the residual is the sum of the district’s real departure from its province and any failure of the offset to transport, 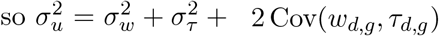 Calibrating the register by (21) does not need 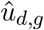 when transport holds and *λ*_*d,g*_ = *λ*_*p*(*d*),*g*_, whereas 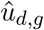 by (24) does not target *η*_*d,g*_ − *η*_*p*(*d*),*g*_ exclusively when transport fails. In Section 4.3 we develop a diagnostic to separate the two terms.

Given (11), the choice between (21) and (25) rests on other considerations. The register variance *v*_*d,g*_ is typically much smaller than 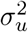 and *ψ*_*p*(*d*),*g*_, so the two estimates of *η*_*d,g*_ are close to each other. In this application they separate by a median of 0.001 to 0.022 on the log-odds scale across the four countries at the age-collapsed fit and 0.002 to 0.012 at the age–sex fit (Appendix B). To estimate *η*_*d,g*_ in a later period with only the register available, both forms hold 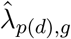 fixed. The calibrated register (21) is then updated by the new *z*_*d,g*_ in full, whereas the shrinkage form (26) is updated by the fraction *γ*_*d,g*_ of it, its survey component *y*_*p*(*d*),*g*_ held at the survey-year value, and the form (25), 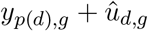, is not updated at all. In a third route, 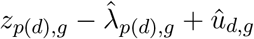, both 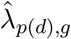 and 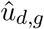 are held fixed and the observed *z*_*d,g*_ for the target cell is ignored. With *γ*_*d,g*_ near one the first two updates differ little, there seems no convincing argument for predicting *u*_*d,g*_ and holding it fixed, and the application reports (21), which uses the current register in full.

### 4.3 A diagnostic of the transport assumption

A transport assumption is necessary for subnational targets, and it warrants examination. Apply the general setup (5) and (6) with *i* = (*d, g*), *η*_*i*_ = logit(*θ*_*i*_) and *δ*_*i*_ = *λ*_*i*_, allowing *λ*_*d,g*_ to differ over *d* ∈ *D*_*p,g*_, and let the survey resolve districts. Then

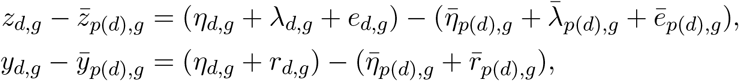

where 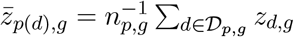 and similarly for the other averages. Differencing the two removes *η*_*d,g*_ altogether and leaves

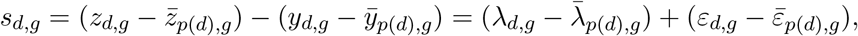

with *ε*_*d,g*_ = *e*_*d,g*_ − *r*_*d,g*_ here, since each district now has its own survey error. The within-province variation of the true positivity drops out of *s*_*d,g*_, so *τ*_*d,g*_ separates from *w*_*d,g*_ in (27). Let

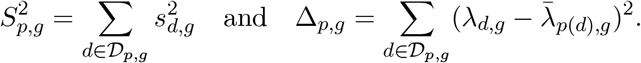

In general 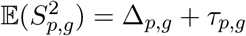, and under (11) Δ_*p,g*_ = 0 and 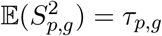, where

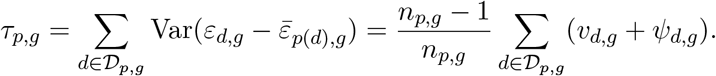

It follows that

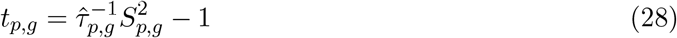

diagnoses the transport assumption. The assumption is plausible where *t*_*p,g*_ is negative or positive and small, and doubtful where it is positive and large. Here *ψ*_*d,g*_ is the design variance of the district survey logit, estimated from the few clusters sampled in the district. Together with *v*_*d,g*_ it determines the noise level 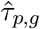. An overstated *ψ*_*d,g*_ pushes *t*_*p,g*_ towards negative values and wrongly favours transport. An understated one pushes *t*_*p,g*_ towards positive values and wrongly suggests a failure. Since 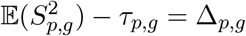, the same quantity estimates the within-province variance of the offset, 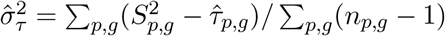, a variance confounded with 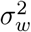 in (27) when the survey resolves provinces alone.

A test needs an assumption about the distribution of *ε*_*d,g*_. The simplest is normality of *e*_*d,g*_ and *r*_*d,g*_, and the second of these is questionable at small cell sample sizes. Remedies are unavailable, especially when resampling methods are not viable without access to all the survey data and design information. Under normality, writing 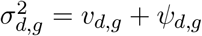,

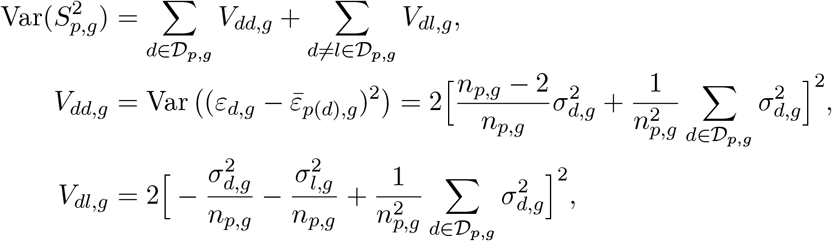

the second line using Var(*X*^2^) = 2 Var(*X*)^2^ and the third Cov(*X*^2^, *Y* ^2^) = 2 Cov(*X, Y*)^2^ for jointly normal variables of mean zero. A test of (11) follows as

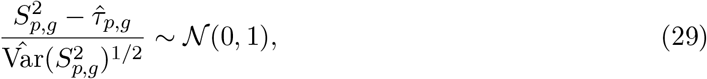

one-sided, since 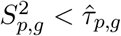 favours (11).

The diagnostic requires survey estimates at district level, which only exist in districts where at least one undiagnosed case was found among the people sampled there, since a district with none has a survey estimate of zero and no logit. A district with only a single cluster sampled is also dropped, because its linearised design variance is exactly zero. Provinces with a single surveyed district contribute nothing. Excluding districts with no undiagnosed case selects on the survey outcome. The excluded districts are those with low survey positivity relative to their register, and hence with large offsets *z*_*d,g*_ − *y*_*d,g*_, so one tail of the offset distribution is removed and 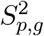 is lowered. The transport assumption therefore tends to be rejected less often, and 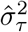 to be understated.

## 5 Application to four countries

### 5.1 Selection offsets, 2020–21

Selection is positive and statistically significant in all but three age–sex groups, all of them women (Figure 1).

**Figure 1.**
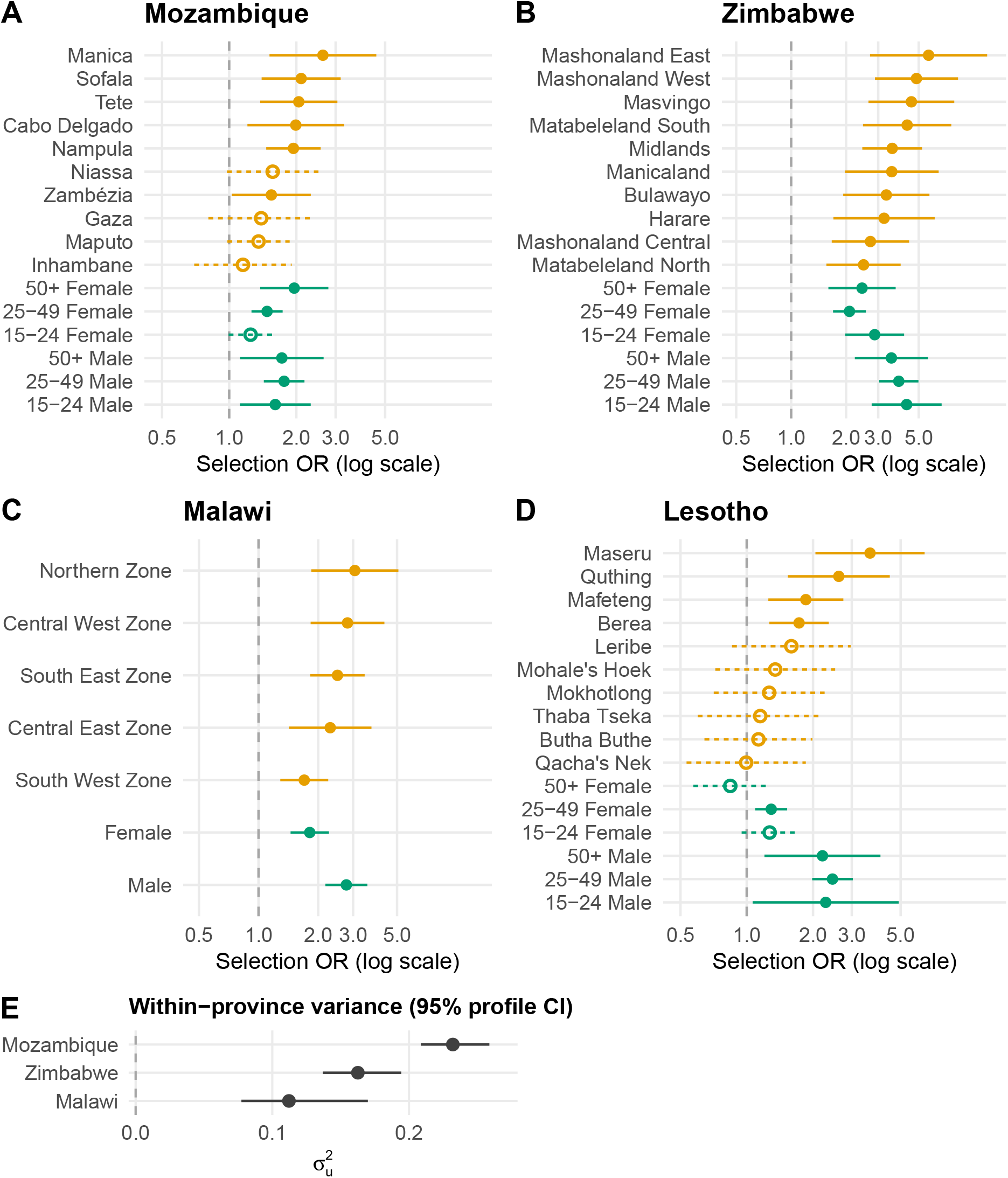
Absolute selection odds ratios in Mozambique (A), Zimbabwe (B), Malawi (C), and Lesotho (D). No additivity is assumed, so the fit does not split selection into a province effect plus an age–sex effect. Provinces are shown in orange and age and sex groups in green, both as margins of the same province-by-stratum cells. The dashed line at 1 means no selection, and filled markers with solid intervals exclude 1, while open markers with dashed intervals include it. Malawi only has two strata, as its routine data are not disaggregated by age. Panel E gives 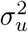 with its 95% profile interval for the three countries with an identified estimate.

The national figure is the average of the province–stratum offsets on the log-odds scale, with each cell weighted by its number of people not yet diagnosed, from the survey’s biomarker weights. Standardised this way the national selection odds ratio is 1.70 (1.50, 1.92) in Mozambique, 3.67 (3.07, 4.38) in Zimbabwe, 2.36 (1.97, 2.83) in Malawi and 1.80 (1.55, 2.08) in Lesotho. In each country an adult who has HIV without knowing it is more likely to present for a routine test than an adult without HIV of the same sex and age in the same district.

Men show stronger selection than women of the same age in nine of the ten comparisons available, consistent with more symptom-driven male testing and fewer routine entry points equivalent to antenatal care (Giguère et al., 2021). Among women in Mozambique, selection rises with age, in line with antenatal testing of younger women regardless of risk (Allorant et al., 2024).

A province summary is the average of its cells over strata on the log-odds scale, with the same stratum weights in every province of a country. Selection is present in all ten provinces of Zimbabwe, at a median odds ratio of 3.57, in six of the ten provinces of Mozambique and in four of the ten districts of Lesotho, and the five zones of Malawi range from 1.70 to 3.06 (Figure 1). Province intervals overlap extensively within every country, so the provinces cannot be ranked by their selection odds ratios.

We also estimate the within-province variance 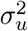 of the area effect *u*_*d,g*_ in (15). The offset captures, for a given province and stratum, the difference between the register-derived and survey-derived estimates. 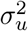 measures the spread of the districts of that province once the offset is removed. Under the null hypothesis 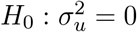, districts of the same province differ only by the sampling noise of the register, of variance *v*_*d,g*_, near 0.01 at the median cell. The survey error does not enter this comparison, because every district of a province is differenced against the same survey estimate *y*_*p,g*_. The inset of Figure 1 shows 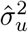 with a 95% profile-likelihood interval, which respects the lower bound of the variance at zero (Stram and Lee, 1994). The estimate is 0.232 (0.209, 0.259) in Mozambique, 0.163 (0.137, 0.194) in Zimbabwe and 0.112 (0.077, 0.170) in Malawi. These values are about twenty times the register noise expected under *H*_0_, which suggests real variation within province–stratum cells. Lesotho is fitted at admin-1, and therefore 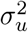 s not identified.

With the diagnostic of Section 4.3, true differences between districts can be separated from a failure of the offset to transport. The diagnostic can be applied in three of the four countries; Lesotho is excluded since analyses are done at the province level. Further, the diagnostic requires that each province has at least two districts with sampled areas; Bulawayo and Harare only have a single district, so only eight of Zimbabwe’s ten provinces are tested. Figure 2 gives the test (29) for the remaining 23 provinces. The test rejects transport at the one-sided 5% level in one province, Mashonaland West in Zimbabwe, about as often as expected by chance across 23 tests, and in none under the reference distribution of Appendix A.5. Table 4 reports the pooled moment estimate 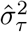 of Section 4.3 for each country. The estimate is 44% of 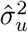 in Mozambique and 64% in Zimbabwe, and negative in Malawi. Because districts with no undiagnosed case are excluded from the diagnostic (Section 4.3), these shares are likely to be understated. At these point estimates the deployment condition of Section 6.2 is close to its threshold in Mozambique and fails in Zimbabwe, but every interval spans zero and the whole of 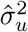, so the evidence on the condition is inconclusive.

**Table 4.** The within-province variance of the offset, 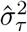 of Section 4.3, by country, pooled over the provinces tested at district level with age and sex collapsed.

| Country | Provinces | Districts | Rejecting | $\hat{\sigma}_\tau^2$ (95% CI) | $\hat{\sigma}_u^2$ |
| --- | --- | --- | --- | --- | --- |
| Mozambique | 10 of 10 | 67 of 128 | 0 | 0.090 (−0.044, 0.223) | 0.202 |
| Zimbabwe | 8 of 10 | 59 of 63 | 1 | 0.080 (−0.128, 0.288) | 0.126 |
| Malawi | 5 of 5 | 25 of 27 | 0 | −0.025 (−0.166, 0.115) | 0.112 |

**Figure 2.**
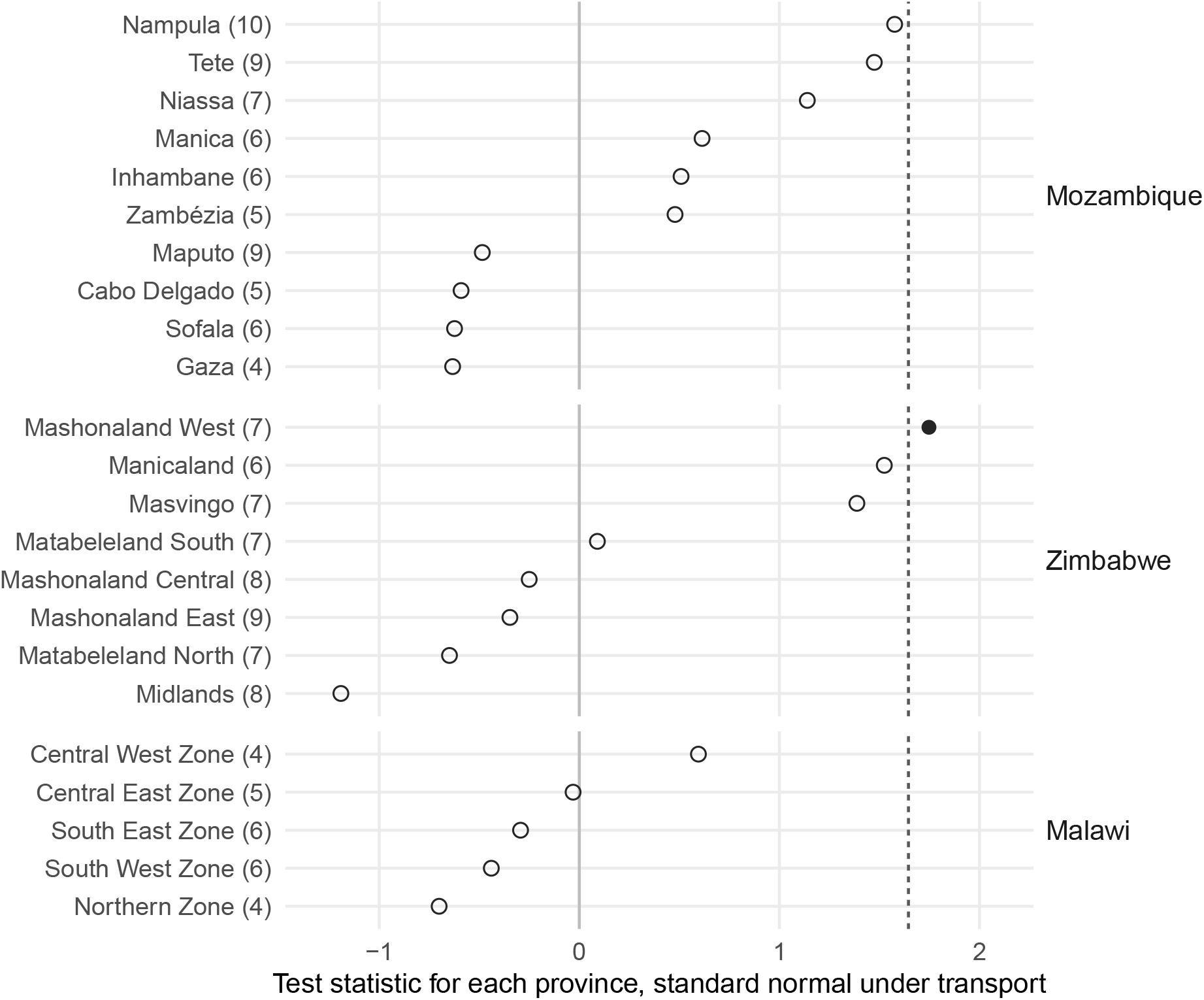
The test (29) of the transport assumption, one point per province, at district level with age and sex collapsed. The number of districts compared is in brackets. Under transport each statistic is referred to a standard normal, and transport is rejected at the one-sided 5% level for values beyond the dashed line, as for the province shown by a filled circle; open circles do not reject. Only districts sampled in more than one cluster and with at least one undiagnosed case are used; Table 4 gives the numbers dropped. Lesotho is excluded by construction.

### 5.2 Register calibration, 2015–16 to 2020–21

Between surveys, the calibrated estimate uses the offset estimated at the last survey, so any drift in the offset since that survey would bias the calibrated estimate. We test for that drift by recomputing the offset at the previous survey round, MPHIA 2015–16, LePHIA 2016–17 and ZIMPHIA 2015–16, for every survey area with routine data at both rounds.

The routine measurement comes from the Monitoring, Evaluation and Reporting testing indicators (HTS_TST and HTS_TST_POS) of the United States President’s Emergency Plan for AIDS Relief (PEPFAR). These keep the same definitions at both rounds, although the testing modalities reported under them were revised in 2018, whereas the indicators and disaggregation of the ministry registers changed between the rounds. PEPFAR records cover only PEPFAR-supported areas, so Lesotho contributes its five western lowland districts, where early support was concentrated. The public data carry no age–sex detail, so the offset is measured at area level only.

Mozambique is excluded, because in IMASIDA 2015 awareness of status was measured from whether a participant had ever tested and received a result, with no awareness question and no antiretroviral assay. MPHIA 2015–16 uses three regions, against the five zones of the 2020–21 round (Table 5).

**Table 5.** Two-round stability of the selection offset. OR_0_, OR_1_ are the mean implied selection odds ratios 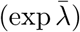 at the 2015–16 and 2020–21 rounds. “Level shift” is 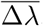, the average round-to-round change in the offset on the log-RTP scale, and “Area drift” is its standard deviation across areas, the part of the movement that is missed when one round’s offset is applied at the next. “Corr” is the Pearson cor(*λ*_0_, *λ*_1_) across areas. “Δ vs prevalence” is cor(Δ*λ*, Δprevalence), with prevalence the biomarker-measured total. The routine source is the PEPFAR MER testing indicators at both rounds, and Lesotho’s five areas are the western lowland districts.

| Country | Areas | $OR_0$ | $OR_1$ | Level shift | Area drift | Corr | $\Delta$ vs prev. |
| --- | --- | --- | --- | --- | --- | --- | --- |
| Zimbabwe | 10 | 2.50 | 3.47 | +0.33 | 0.31 | 0.21 | −0.51 |
| Malawi | 3 | 2.54 | 3.06 | +0.18 | 0.07 | 0.83 | −0.47 |
| Lesotho | 5 | 1.00 | 1.51 | +0.41 | 0.25 | 0.81 | −0.88 |

The offset rises between the rounds in all three countries (Table 5), by +0.33 on the log-odds scale in Zimbabwe, from an implied selection odds ratio of 2.50 to 3.47, by +0.18 in Malawi and by +0.41 in Lesotho. Within Zimbabwe, where all ten admin-1 areas are available at both rounds, the areas move by different amounts and change their relative positions, with a round-to-round correlation of only 0.21. In Zimbabwe, the share of the across-area variance of the offset due to sampling error is 75% at round 0 and 49% at round 1. Even if the areas kept their relative positions exactly, the correlation could therefore not exceed about 0.36. The low value is at least as much a consequence of this imprecision as of any change in the areas’ relative positions. Each round’s offset is estimated with sampling error, so we test each area’s change against no movement, dividing it by the square root of the sum of the two rounds’ sampling variances. At the 5% level the test rejects *H*_0_ in six of the eighteen admin-1 areas across the three countries, so some of the movement exceeds sampling noise. The area drift of Table 5 includes sampling error at both rounds. Net of that error, the area drift is 0.18 in Zimbabwe, 0.14 in Lesotho and indistinguishable from zero in Malawi.

The drift of the offset between rounds seems negatively correlated with the change in the area prevalence; −0.51 in Zimbabwe, − 0.47 in Malawi and −0.88 in Lesotho. This is consistent with selection rising where the epidemic receded faster, and with the shift of PEPFAR-supported programmes toward targeted, higher-yield testing between the two rounds.

Part of that correlation is mechanical, because *λ* is differenced against the same survey that measures prevalence, so the two share their sampling error. The mechanical values, derived from the design variances, are −0.23 in Zimbabwe, −0.22 in Lesotho, and −0.61 in Malawi. The observed correlations exceed them in magnitude in Lesotho and Zimbabwe, though with five and ten areas the difference is not statistically distinguishable, and in Malawi the observed correlation is weaker than the mechanical one.

The round-1 offsets measured here differ from those of Section 5.1, lower in Zimbabwe (3.47 against 3.67) and Lesotho (1.51 against 1.80) and higher in Malawi (3.06 against 2.36). Awareness is measured the same way on both sides at round 1, so the differences are due to PEPFAR-supported testing against the ministry register, the different sets of areas, the collapsing of age and sex, sampling error, and the unweighted areal means used here.

With two rounds of survey and register data, we can also test the estimates available to a programme in 2020–21 in the absence of the 2020–21 survey. Three approaches would be available: (a) reporting the register’s logit test positivity *z*_*d,g*_ for 2020–21 and ignoring the 2015–16 survey data altogether (“Register alone”); (b) combining register and survey to estimate 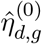 for 2015–16 directly and reusing that estimate for 2020–21 (“Freeze”); or (c) applying the 2015–16 offset to the live register as 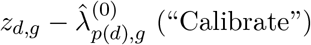. For each area, we derive round 1 estimates using all three (Table 6).

**Table 6.** Three approaches to 2020–21 undiagnosed positivity using the 2015–16 survey and the 2020–21 register. Entries are the root-mean-squared error of undiagnosed positivity in percentage points across the areas of Table 5, against the round-1 survey estimate. “Register alone” applies no calibration to the round-1 register, “Freeze” reuses the round-0 estimate of the target, and “Calibrate” applies the round-0 offset to the round-1 register.

| Country | Areas | Gap (years) | Register alone | Freeze | Calibrate |
| --- | --- | --- | --- | --- | --- |
| Zimbabwe | 10 | 5 | 4.35 | 1.68 | 0.94 |
| Malawi | 3 | 5 | 2.00 | 1.43 | 0.24 |
| Lesotho | 5 | 4 | 1.51 | 2.62 | 1.44 |

Calibrating is the most accurate of the three approaches in all three countries. It improves on freezing by 0.74 to 1.19 percentage points. The register alone is the worst of the three in Zimbabwe and Malawi, by 3.41 and 1.76 points against the calibrated estimate. Under freezing, the current period data are not used, and in Lesotho freezing is less accurate than leaving the register uncalibrated. In Lesotho the calibrated estimate’s margin over the raw register narrows to 0.07 points. There the round-0 offset averages zero (an odds ratio of 1.00) and is negative in three of the five areas, so calibrating with it does not remove the round-1 selection. The calibrated estimate’s bias is larger than the raw register’s (1.24 against 1.11 points), and its root-mean-squared error is smaller only because its variance across areas is lower.

Undiagnosed positivity fell in all eighteen areas and the offset rose in seventeen, so an offset held at its round-0 value is too small and the calibrated register estimate is above the target. Its bias is below both the uncalibrated register’s and the frozen estimate’s in Zimbabwe and Malawi, and above the uncalibrated register’s in Lesotho. Where undiagnosed positivity rises, the relative test propensity would be expected to fall and the calibrated register estimate to be below the target. As long as the offset remains positive, the target is then between the calibrated and the uncalibrated register. With three countries and one interval between rounds, neither direction can be established in general. This ordering concerns the bias alone. It implies nothing about the root-mean-squared error, which also depends on the size of the offset’s drift.

From the two-round comparison we therefore find that the offset moved in these data, and we estimate the size of that movement with large uncertainty.

## 6 Simulation study: conditions for calibration to improve on current practice

In the four-country application we estimate the offset and its uncertainty, but an external benchmark would be needed to assess the accuracy of these estimates. We therefore generate a synthetic population of districts observed over several annual waves, in which the same estimand is recorded in a register and measured in a probability audit, as in the two equations of Section 3.3, and the truth is known. We compare the calibrated estimate with four forms of current practice: the survey alone (with the stratum mean of the audited districts in each unaudited cell), the uncorrected register, the last survey estimate held fixed between surveys, and one offset for all age–sex strata. Below we describe the simulation setup in Section 6.1, and investigate the conditions for successful statistical calibration in Sections 6.2 to 6.4. Table 8 summarises the results and findings.

### 6.1 A temporal generative model

The population is a country of *P* = 10 provinces with four districts each, so *D* = 40 districts, observed over *T* = 6 annual waves. The estimation unit is a district by age–sex cell, with strata for women and men aged 15–24 and 25–49. These dimensions are within the range present across the four countries of Section 5, which run from 10 to 159 areas nested in five to ten provinces.

For cell *i* = (*d, g*) in province *p* at wave *t* = 0, …, *T* − 1, true prevalence *q*_*it*_ and awareness *a*_*it*_ are generated on the logit scale and combined into the estimand of Section 2, the positivity among people not yet diagnosed, *θ*_*it*_, with working-scale value *η*_*it*_ = logit *θ*_*it*_:

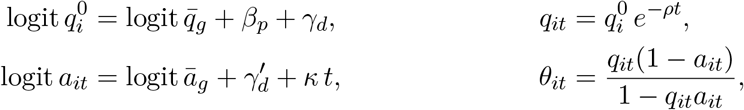

with stratum baselines 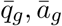 set to values consistent with PHIA estimates, province effects *β*_*p*_, district effects 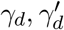, prevalence declining at rate *ρ* on the probability scale, and awareness gaining *κ* per wave on the logit. The selection offset has a stratum baseline that is larger for men, who are tested more selectively, and a district term. The district standard deviations of prevalence, awareness and the offset are scaled by a common factor so that the within-province variance 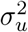 matches 0.20, between the main-fit values of Zimbabwe (0.16) and Mozambique (0.23), with 34% of it due to transport failure of the offset. Appendix C gives every parameter value and the resulting epidemiology.

The offset’s own dynamics are set as four scenarios. Under the constant scenario the offset does not move, so the calibration derived from one survey remains valid throughout the period. Under drift, the offset of each unit changes by a constant amount at every wave, and that amount is drawn separately for each unit. Under the shock scenario, a break affecting all areas happens at wave 3 and persists, as a stylised interruption to testing services due to funding or the arrival of a new testing modality, such as self-tests. Under the reshuffle scenario, we use a stationary first-order autoregression in each unit, so areas change their relative order between waves. Figure S2 in Appendix C shows the paths of *λ* and *η* under all four scenarios. In the generative model *η* is built from prevalence and awareness before the offset dynamics are applied, so no scenario is more favourable to the estimand than another.

On the working scale, the register records *z*_*it*_ = *η*_*it*_ + *λ*_*it*_ + *ε*_*it*_ with *ε*_*it*_ ~ *N*(0, *v*), and the audit measures 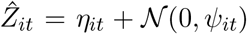 in the districts surveyed at wave *t*, as in equations (12) and (13). The register variance *v* is small, while *ψ*_*it*_ is the design variance of 120 adults per stratum in each audited district at a design effect of 2. Eight of the forty districts are audited at each wave, drawn uniformly at random, so each wave’s audit has 3,840 adults, about a quarter of a PHIA round. The calibrated estimate is the register estimator (21), 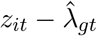, in every cell, with 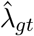 estimated from the audited districts of stratum *g* at wave *t*. The survey-only estimate is the direct estimate in audited cells and, elsewhere, the mean of the audited direct estimates in the same stratum and wave. Appendix C gives both estimators in full. This survey-only estimate is computed without auxiliary data, so each margin reported here is the reduction in root-mean-squared error obtained by adding the register to the audit.

### 6.2 Reporting level and transport failure

Under each of the four scenarios, the calibrated estimate has the lowest root-mean-squared error of the three estimators at every reporting level, from national down to the district by age–sex cell (Table 7). Its margin over the audit is smallest at national level, at 0.26 to 0.35 percentage points across the scenarios, and between 1.37 and 1.89 below it. At national level the survey is at its design resolution, and below it a probability survey is noisier while the register keeps its coverage. The register alone is the least accurate at every level. The 95% intervals of the calibrated estimator have empirical coverage between 0.895 and 0.908 across the scenarios, with median widths of 4.0 to 4.9 percentage points (Appendix C).

**Table 7.**
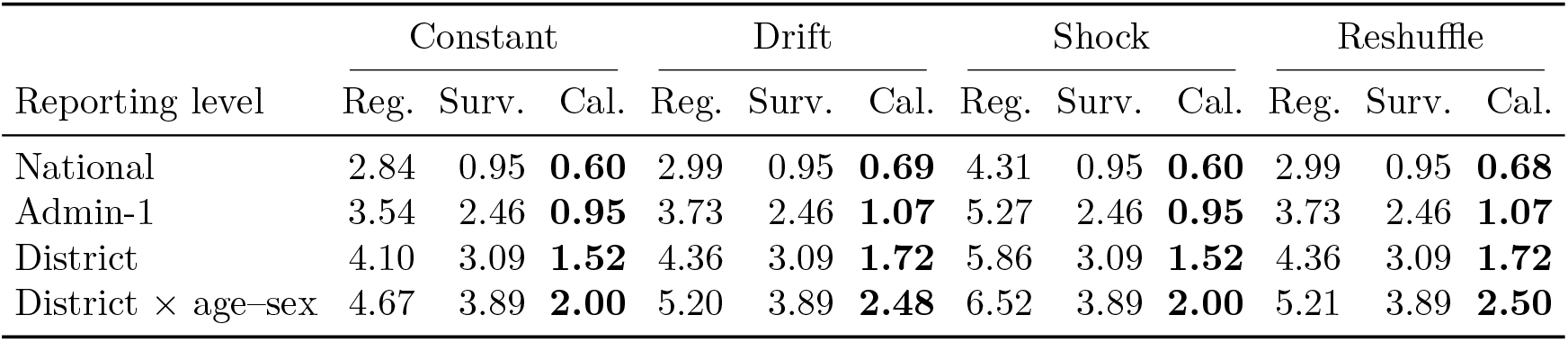
Root-mean-squared error of estimated undiagnosed positivity, in percentage points, by reporting resolution under each offset scenario of Section 6.1, for the register alone (Reg.), the survey audit alone (Surv.) and the calibrated estimate (Cal.). The survey-only error is the same under every scenario, and the calibrated error under a shock equals that under a constant offset, because the offset is re-estimated at each wave. In bold, the lowest error in each scenario and row.

For an area outside the survey sample, the alternative to the calibrated register 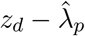 is the province survey mean applied to every area. Under model (15), the calibrated register has the smaller mean squared error against the area truth when

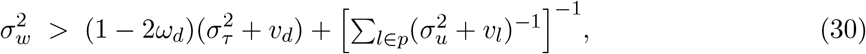

where the sum is over the areas *l* of province *p* that contribute to 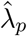 and *ω*_*d*_ is the weight of area *d* in (19). This follows from the two errors against the area truth, 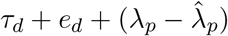 for the calibrated register and *r*_*p*_ − *w*_*d*_ for the province mean, with Cov(*w*_*d*_, *τ*_*d*_) = 0, so that 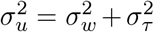 (Section 4.2). Area is one of the areas from which 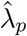 is estimated, so the two parts of the calibrated error are correlated, with covariance 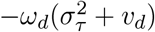. Taking variances and substituting (20) for Var 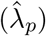, the survey’s own error *ψ*_*p*_ appears on both sides and cancels, which gives (30). With equal register variances *v, ω*_*d*_ = 1*/m* for *m* areas and the bracketed term is 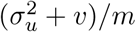 and the condition reduces to 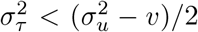 for any number of areas. Calibrating an area outside the survey sample therefore lowers its prediction error approximately when less than half of the within-province variance is due to failure of the transport assumption, and raises it otherwise. The two components can be separated with the diagnostic of Section 4.3 where the survey resolves districts. In the simulation, the calibrated estimate remains more accurate than the audit at every reporting level as 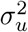 varies from 0.05 to 0.50, and at every subnational level as the share due to transport failure varies from 20 to 80% (Appendix C).

### 6.3 The offset’s drift between surveys

Between surveys, two procedures are available: freezing and updating. Under freezing, the calibrated estimate from the last survey is reused. Under updating, the offset estimated at the last survey is applied to the register of the current year. With the survey alone, freezing is the only option. Over one year, the error of the frozen estimate is the change in the target, Δ*η*, and the error of the updated estimate is the change in the offset, Δ*λ*. To a first approximation, the one-step-ahead mean squared errors on the logit scale are

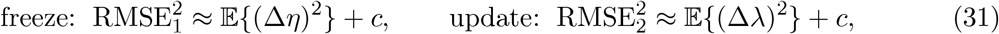

where *c* denotes the calibration-precision terms common to both. Each mean squared change is the square of the change common to all areas plus the variance of the change across areas.

Applying the last survey’s offset to the current register is therefore more accurate than reusing the last calibrated estimate whenever the offset changes less than the target between surveys, that is, whenever *r* = E {(Δ*λ*)^2^ */*E (Δ*η*)^2^} *<* 1. In the two-round data of Section 5.2, *r* = 0.41 across the three countries measured on one awareness definition, and the upper end of its 95% interval is 0.81.

The approximation holds only if *c* does not change with the drift. In the simulation, each unit’s offset drifts at its own rate, so the spread of the offset across areas grows, and *c* with it. We therefore locate the crossing point by simulation, with the annual drift expressed as a fraction of the offset’s spread across areas within one year (Appendix C). With no drift, updating is 0.39 percentage points more accurate than freezing, and beyond a drift of 0.44 times the spread freezing is more accurate.

Under a shock, the offset moves in one step by an amount unforeseeable from the last survey, and the calibrated estimate keeps that bias until the offset is re-estimated from the next survey. Nothing in the model protects against this; the only protection would be frequent audit surveys.

### 6.4 A register without age and sex detail

If a register does not record age and sex, a single offset can be estimated for each area. That offset is more precise, since it is estimated from all strata at once, and biased for every stratum whose selection departs from the area average. Pooling is therefore more accurate than stratum-resolved fitting when the saving in variance exceeds the added squared bias. That bias grows with the between-stratum spread, the standard deviation of the offset across the strata of an area on the log-odds scale. A spread of zero means selection is identical in every stratum and pooling costs nothing, and a large spread means separate offsets are needed for the strata. The two costs balance at a spread of about 0.30. Mozambique (0.16) and Zimbabwe (0.28) lie below that point, Malawi (0.30) at it and Lesotho (0.43) above it, so pooling gains 0.23 and 0.04 percentage points in Mozambique and Zimbabwe, is neutral in Malawi and costs 0.32 in Lesotho (Table S10 in Appendix C). At every spread, the pooled calibration is also more accurate than either source used alone.

## 7 Discussion and conclusion

We formalise a statistical framework that combines a probability survey with a routine health information system, for estimating service coverage and disease burden below the national level. Our four main contributions are to: (a) identify and model the register’s bias wherever the two sources overlap; (b) apply that model to the live register, so a programme reports estimates even in years without a survey; (c) use a second survey round to measure how far the bias moves between rounds; and (d) build a known-truth simulation that fixes the conditions under which the calibrated framework improves on current practice (Table 8).

**Table 8.** Key results and conclusions of the simulation study, by subsection, with the evidence from the four countries. Reductions in error are in root-mean-squared error, relative to the alternative.

|  |  |
| --- | --- |
| **6.2 Reporting level and transport failure** |  |
| Below the survey's resolution | Under every offset scenario, the calibrated estimate has the lowest root-mean-squared error of the three estimators at every reporting level. Its error is 27 to 37% below the survey's at national level and 36 to 61% below it at subnational levels, and the register alone is the least accurate everywhere. |
| Outside the survey sample | Calibration lowers the prediction error of an unsampled area approximately when less than half of $\sigma_u^2$ is due to transport failure of the offset, condition (30). In the simulation the reduction narrows as that share rises but stays positive against the survey alone. |
| Four countries | PHIA surveys are designed for admin-1 estimates, and estimates are needed by district, age and sex. The transport-failure share is 44% in Mozambique, 64% in Zimbabwe and below zero in Malawi, each with an interval spanning zero to one. |
| Conclusion | Wherever estimates are needed below the survey's design resolution, calibration improves on both sources. Whether it also improves on the province survey mean in an unsampled area depends on the transport-failure share, and the evidence on that share is inconclusive. |
| **6.3 The offset's drift between surveys** |  |
| Freezing or updating | Applying the last survey's offset to the current register is more accurate than reusing the last calibrated estimate whenever the offset changes less than the target between surveys, $r < 1$. In the simulation, updating stays more accurate up to an annual drift of 0.44 times the offset's spread across areas. A shock to the offset is corrected only by the next audit. |
| Four countries | $r = 0.41$ (upper end of the 95% interval 0.81) across three countries, and over the real gap between rounds applying the last offset to the live register was 0.74 to 1.19 points more accurate than freezing (Table 6). |
| Conclusion | Between surveys, the last survey's offset should be applied to the live register, and only frequent audits protect against a shock. |
| **6.4 A register without age and sex detail** |  |
| One offset per area | One offset for all strata is more accurate than stratum-resolved offsets when the offset's spread across the strata of an area is below 0.30 on the log-odds scale, and at every spread the pooled calibration is more accurate than either source used alone. |
| Four countries | The spread is 0.16, 0.28, 0.30 and 0.43 in Mozambique, Zimbabwe, Malawi and Lesotho, so pooling is more accurate in the first two, equally accurate in Malawi and less accurate in Lesotho. |
| Conclusion | A register without age and sex detail can still be calibrated, and where selection differs little between strata a single offset per area is the more accurate choice. |

### 7.1 Relation to existing methods

The bias between the two sources reduces to a single additive term on the log-odds scale, identified wherever the survey and the register overlap. In these four countries, it is large, systematic across age–sex strata and far larger than sampling noise. Subtracting the survey from the register wherever both exist gives an unbiased direct estimate of the offset in that cell. Every gain therefore comes from the structure placed on the offset across cells, and from the transport assumption (11) behind that structure.

Within HIV surveillance, this arrangement has a direct predecessor. From the mid-2000s, national HIV estimates in high-burden African countries were based on sentinel serosurveys at a panel of antenatal clinics, calibrated against HIV prevalence measured in national population-based surveys (UNAIDS and WHO, 2003; Ghys et al., 2004; Gouws et al., 2008; Marsh et al., 2014). In that calibration, one bias term was estimated for each epidemic subpopulation and held constant over time (Marsh et al., 2014). Here the offset is indexed by district and age–sex stratum and transported to unsampled areas. Once routine testing of pregnant women became near-universal, sentinel serosurveys were replaced by routine testing records from antenatal care in most countries (WHO, 2015; Sheng et al., 2017, 2020; Maheu-Giroux et al., 2019a). The framework here adds no purpose-built collection, because the register already exists and the audit is a smaller version of existing household surveys.

In official statistics, the closest precedent is census coverage evaluation. A post-enumeration survey independently re-measures a probability sample of areas, and the enumeration’s error is estimated by dual-system estimation (e.g. Hogan, 1992). Offices moving to combined and register-based censuses face the problem of a drifting offset, since the coverage error estimated at one census becomes outdated over the intercensal period (UNSD, 2025). Guidance for those audits is being written for the next census round (UNFPA, 2026).

Across the three countries where the offset could be compared between rounds, the estimated offset shifted upward. In two of the three, the correlation of the change in the offset with the change in area prevalence exceeded in magnitude the value implied by shared sampling error. With five and ten areas, neither difference was statistically significant. The bias of an estimate that holds the last survey’s offset fixed had the same sign in all three countries. In all three countries, applying the last survey’s offset to the live register over the real gap between rounds lowered the root-mean-squared error against reusing the last round’s estimate. Comparisons of antenatal clinic and population prevalence across several survey rounds (Eaton et al., 2014), re-expressed on the offset’s log-odds scale, could give an external estimate of how fast a selection factor changes between surveys.

The calibration lowers the root-mean-squared error against either of the data sources used alone at every reporting level, and Section 6 fixes the conditions under which that holds. Using the register alone to derive estimates is the worst of the three approaches at every level. In that regard, our approach differs from the literature on non-probability samples. In that literature, the outcome is measured in the non-probability source alone, and either the probability that a person is included in it is modelled or estimates are calibrated on auxiliary variables shared with a probability sample (Elliott and Valliant, 2017; Chen, Li and Wu, 2020; Beaumont, 2020; Kim and Tam, 2021; Sen and Lahiri, 2026). Our setting differs in two ways. The outcome is measured in both sources, so the bias is identified directly wherever they overlap. And the non-probability source is large, so its benefit is spatial reach more than precision. The register provides an estimate for every district, and under the transport assumption most of the district prediction variance comes from the calibration term (Appendix B). When failure of that assumption is allowed for, most of it comes from the transport-failure term.

### 7.2 Limitations

Our study has several limitations. First, our model rests upon the transport assumption (11), under which the selection offset is the same in every district of a province and stratum. With district-level covariates, this can be relaxed so that 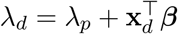 varies within province. The district residual of the fitted model combines the district’s true departure from its province with any failure of the offset to transport. The two are confounded when the survey is analysed at province level. However, when the survey is analysed at district level, the split between the two is identified from one round, using the diagnostic proposed in Section 4.3. In the three countries with a usable diagnostic, the test rejects transport in a single province, about as often as expected by chance. The confidence intervals on the within-province variance of the offset, 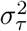, are too wide to identify the dominant component (Table 4). The district intervals therefore include the estimate 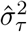, which widens the median interval from 2.7 to 5.0 percentage points in Mozambique and from 1.7 to 2.8 in Zimbabwe (Table S5). That estimate is itself imprecise, so these intervals are approximate, and in Lesotho, where no diagnostic is possible, they are conditional on transport.

Second, the fitted offsets are upper bounds on selection wherever re-testing by people already diagnosed goes uncorrected, since a test on someone already diagnosed is positive and routine positivity is inflated by such tests. A share of re-tests common to a country cannot exceed the positivity of its least positive area. Removing a share of that size from the national register positivity lowers its log-odds by 0.28 in Mozambique, 0.27 in Zimbabwe, 0.37 in Malawi and 0.28 in Lesotho, and the national selection odds ratios become 1.29, 2.79, 1.64 and 1.36, so positive and material selection remains in all four countries. The bound is a national one, since at the least positive area itself the corrected positivity would be zero.

Third, the two sources measure different things in time. The survey measures the stock of people not yet diagnosed at fieldwork, whereas the register accumulates positive tests over a year, during which incident infections enter that pool and newly diagnosed people leave it. Incident infections that are tested within the year appear in the register but not in the survey’s stock, so incidence raises register positivity relative to the stock and is absorbed into *λ*, in proportion to the ratio of annual incidence to undiagnosed positivity. Diagnoses made during the year deplete the stock, and if testing reaches the most likely positives first, the register positivity of later months targets a less positive pool. We align the register to the calendar year of fieldwork, and a twelve-month window centred on the fieldwork is the natural sensitivity analysis. Such misalignment is unavoidable as long as the survey is not continuous over time, and it is accepted in official statistics outside the few surveys fielded continuously, such as labour force surveys.

Fourth, the register is indexed by the district of the facility and the survey by the district of residence. Care sought outside the district of residence, at urban and referral facilities and across district boundaries, assigns tests to the wrong district. Within a province this contributes to the district departure 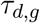 and leaves 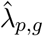 unaffected, whereas flows across a province boundary bias the province offset itself.

Fifth, the framework inherits the register’s data quality. Zimbabwe’s extract reports Harare and Bulawayo as health units of which seven, central hospitals among them, record between 575 and 23,638 tests and no positive, while the other units of the two cities run at 5 to 26 per cent positivity. Treated as districts, those seven units inflated Zimbabwe’s 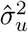 by about 45% and moved the city offsets, Harare’s from 3.23 to 4.25 and Bulawayo’s from 3.32 to 3.05. We pool each city to one area, which leaves the province totals unchanged. Checks of this kind, for units with implausible positivity or with tests recorded away from their positives, belong to the routine quality assessments that national systems now run.

### 7.3 Future work

Calibrating a district outside the survey sample lowers its prediction error approximately when 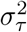 is below half of 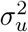 (Section 6.2). The point estimates of Section 5.1 lie near that threshold in Mozambique and above it in Zimbabwe, and the intervals are too wide for either to be established. The width of the confidence interval on 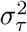 indicates the gain in district-level survey precision required to tell on which side of that threshold 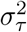 lies. It is also a concrete target for the design of a future survey audit. The next survey audit should ideally be designed so that this interval lies entirely below or entirely above that threshold. In such an audit, fewer areas would be visited, deeply enough to estimate the area truth directly. With an audit spread thinly to estimate province levels, 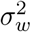 and 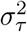 stay confounded, and by (20) the variance of 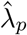 cannot fall below the survey term *ψ*_*p*_ however many districts are added. Where a survey audit of a given size is placed is a design question of its own. Each round’s probability sample could be allocated according to the register–survey discrepancy, so that subnational accuracy is maintained at a fixed budget as the offset moves. Graphical and algorithmic methods for constructing designs that optimise a chosen criterion (Panahbehagh, 2026) are a natural tool for that allocation. With a continuously fielded survey (Roux, 2020), the offset could instead be re-estimated over a sliding window.

Between surveys, the offset is held at its last estimate. It could instead be updated from district covariates that the routine system records every year, with the relation between the offset and those covariates estimated at one survey round and checked at the next, as in Section 5.2. Statistical calibration in general combines two measurements of one quantity, one precise and costly, the other imprecise and cheap, so the framework is not limited to the setting here. Directly applying it beyond HIV requires a routine source that is close to complete for its own population, a probability sample measuring the same quantity on an overlapping frame, and a scale on which the bias between them is additive. The additivity used here follows from the derivation of Section 3, in which the ratio of test propensities for people with and without HIV enters the log-odds of routine positivity as a single term. The same additivity holds for another indicator where entry into the routine record also takes the form of a ratio of propensities between the group counted and its complement, as for malaria test positivity in routine testing against parasite prevalence in a household survey, or for tuberculosis notification. Where entry depends on the outcome in some other way, the link *h* of Section 3.1 is left free. For childhood immunisation, administrative coverage is computed as recorded doses over an estimated target population. Errors in either term are multiplicative, so the bias is additive on the log scale. Where no link makes the offset additive, the framework does not apply.

Statistical calibration is the middle stage of a trajectory for register-based inference. At one end is data integration, in which the survey remains indispensable. At the other is targeted audit sampling, in which the register is trusted and is only checked and adjusted with a lightweight survey. Health ministries will move along this trajectory as the quality of routine health information systems continues to grow and funding for large-scale household surveys decreases.

### 7.4 Conclusion

Routine health information systems have spread across the world, and their quality is improving, helped by the spread of automated data checks. A register of service contacts, calibrated by a small, well-placed probability sample, can be a tremendous source of subnational estimates that can guide local health policy making. These estimates come from data owned by the country, collected in the course of service delivery, and available throughout the year. In the meantime, funding for large household surveys is declining. The growing availability of such large routine data sources is to be welcomed, especially where they can be paired with the traditional device of a randomised sample to correct their selection bias.

Statistical calibration does not always reduce the error. It is useful where estimates are needed, to orient policy and interventions, at a resolution for which surveys are not powered, and where reducing the register’s selection bias outweighs the variance added by the correction. Where those conditions hold, a robust and internally consistent routine health information system, calibrated by small targeted audits, is a country-owned, timely and financially sustainable basis for disease surveillance and the estimation of service coverage. It is also a natural stepping stone to the more cost-effective situation in which the register’s bias is small enough for its uses, so that a lightweight validation audit suffices from time to time.

## Acknowledgements

The authors thank colleagues at the University of Southampton and at the participating ministries of health for helpful discussions, and the PHIA Project and the national health information system custodians for facilitating access to the data.

## Data availability

The PHIA survey microdata are available to registered users from the PHIA Project data portal (https://phia-data.icap.columbia.edu/) under its data-use agreement. The routine HIV testing data are the property of the national ministries of health and cannot be redistributed; the routine counts aggregated by country, year, age group and sex, the model outputs behind every table and figure, and the analysis code are available at https://github.com/aallorant/rhis-calibration. The PEPFAR Monitoring, Evaluation and Reporting (MER) testing indicators used in the two-round comparison were obtained from the amfAR PEPFAR data portal (https://mer.amfar.org/).

## Ethics

This study is a secondary analysis of de-identified survey microdata and of aggregate, non-identifiable routine health information system data. The PHIA surveys were approved by the relevant national ethics committees and institutional review boards in each participating country, and the PHIA microdata are released in de-identified form under the PHIA data-use agreement; the routine data analysed here are aggregate counts that contain no personal information. The present secondary analysis of de-identified and aggregate data did not involve contact with human participants and required no separate ethics approval.

