## Supplementary Material for "A statistical calibration framework for subnational health surveillance from routine data and survey audits"

#### A Estimation details

##### A.1 GLS

Every district in a province is differenced against the same survey estimate $y_{p,g}$, so the sampling errors are correlated within a province and the working covariance (18) is not diagonal. Standard small-area software assumes a diagonal covariance. Below we show that the shared term leaves the offset and the shrinkage weight unchanged and enters only the variance of the offset.

Write $A_{p,g} = \sigma_u^2 I_{n_{p,g}} + \mathrm{diag}(v_{d,g})_{d\in p}$ for the covariance without the shared term, and $a_{p,g} = \mathbf{1}^\top A_{p,g}^{-1}\mathbf{1}$, so that $V_{p,g} = A_{p,g} + \psi_{p,g}\mathbf{1}\mathbf{1}^\top$. The Sherman–Morrison identity gives $\mathbf{1}^\top V_{p,g}^{-1} = (1+\psi_{p,g}a_{p,g})^{-1}\,\mathbf{1}^\top A_{p,g}^{-1}$, and that scalar cancels between the numerator and the denominator of (19). The offset is therefore the generalised least squares estimator under $A_{p,g}$ alone, so $\psi_{p,g}$ drops out of the weights. The realised survey error still shifts $\hat\lambda_{p,g}$ as a whole, and it appears in the variance below. Contracting the same identity with $\mathbf{1}$ gives $\mathbf{1}^\top V_{p,g}^{-1}\mathbf{1} = a_{p,g}/(1+\psi_{p,g}a_{p,g})$, whose reciprocal is $\psi_{p,g} + a_{p,g}^{-1}$. Since $A_{p,g}$ is diagonal, $a_{p,g} = \sum_{d\in p}(\sigma_u^2 + v_{d,g})^{-1}$. That is (20). The shared survey error appears there as a term that does not fall as districts are added.

The same cancellation applies to the shrinkage weight. Generalised least squares makes the residual $\tilde\delta_{p,g} - \hat\lambda_{p,g}\mathbf{1}$ orthogonal to $\mathbf{1}$ under $V_{p,g}^{-1}$, and therefore under $A_{p,g}^{-1}$ as well, so the rank-one term annihilates that residual and the district effect is predicted against $A_{p,g}$ alone. The weight is $\gamma_{d,g} = \sigma_u^2/(\sigma_u^2 + v_{d,g})$, and $\psi_{p,g}$ does not enter it.

One term is missing from the prediction variance, and its sign is known. Equation (22) treats $\hat\lambda_{p(d),g}$ as independent of $e_{d,g}$, although district $d$ enters the precision-weighted mean (19) from which the province offset is formed. The omitted term is $2\,\mathrm{Cov}(e_{d,g}, \hat\lambda_{p,g}) = 2\,\omega_{d,g}v_{d,g}$, where $\omega_{d,g} = (\sigma_u^2 + v_{d,g})^{-1}/a_{p,g}$ is the weight of district $d$ in (19). Where $v_{d,g} \ll \sigma_u^2$ those weights are close to $1/n_{p,g}$, so the term is of order $v_{d,g}/n_{p,g}$ and negligible beside (22). It enters negatively, so the reported intervals err wide. At the age–sex cell the weights are far from uniform and that order argument no longer applies.

The derivation excludes one case, which is why Lesotho differs in every comparison below. Where a province contributes a single area, $\hat\lambda_{p,g} = \tilde\delta_{d,g} = z_{d,g} - y_{p,g}$ exactly, so the calibrated register $z_{d,g} - \hat\lambda_{p,g}$ equals $y_{p,g}$ and (26) returns the survey estimate for any value of $\gamma_{d,g}$. The register contributes nothing to the prediction and $\sigma_u^2$ is unidentified. The offset itself is an unbiased raw difference whatever the group size, so such a fit still supports the offset estimates of Section 5, and no district prediction beyond the survey's own. Lesotho's age–sex fit is fitted at admin-1 with one unit per province and stratum, so it is saturated in this way throughout.

##### A.2 TMB

In the latent-field route, the two observation equations (12) and (13) are fitted as a single joint likelihood, with the latent field $\{\eta_{d,g}\}$ treated as a random vector. With the offsets $\lambda$ and the province means $\eta_{p,g}$ as fixed effects, and within-province scale $\sigma_u^2$, the joint negative log-likelihood is

$$\begin{aligned}\ell(\lambda, \sigma_u^2, \eta \mid \text{data}) = &\sum_{(p,g)\in\mathcal{S}} \tfrac{1}{2}\left[\frac{(\hat{\eta}^S_{p,g} - \eta_{p,g})^2}{\psi_{p,g}} + \log(2\pi\psi_{p,g})\right] \\ &+ \sum_{(d,g)} \tfrac{1}{2}\left[\frac{\left(z_{d,g} - \eta_{d,g} - \lambda_{p(d),g}\right)^2}{v_{d,g}} + \log(2\pi v_{d,g})\right] \\ &+ \sum_{(d,g)} \tfrac{1}{2}\left[\frac{\left(\eta_{d,g} - \eta_{p(d),g}\right)^2}{\sigma_u^2} + \log(2\pi\sigma_u^2)\right]. \end{aligned} \tag{S1}$$

The field $\eta$ is integrated out by Laplace approximation (Tierney and Kadane, 1986). For each candidate $(\lambda, \sigma_u)$, the conditional mode is found, $\eta \mid$ data is approximated as Gaussian at the mode with the Hessian of $\ell$ as precision, and the result is substituted back to give a marginal $\ell^\star(\lambda, \sigma_u)$. That marginal is minimised by quasi-Newton, with gradient and sparse Hessian obtained by automatic differentiation (Kristensen et al., 2016; Thorson and Kristensen, 2016; Osgood-Zimmerman and Wakefield, 2023). The procedure is empirical Bayes in the frequentist sense. The latent-field penalty is not a prior, $\lambda$ and $\sigma_u^2$ have no priors and are estimated by maximum marginal likelihood, and the district quantities are conditional modes with delta-method standard errors.

##### A.3 INLA

The integrated nested Laplace approximation (Rue et al., 2009) is an alternative to Markov chain Monte Carlo sampling for latent Gaussian models. A model of that class combines observations that are conditionally independent given a latent field, a Gaussian field with a sparse precision matrix, and a small number of hyperparameters. The model (15) is of that form. The offsets and the field enter linearly, the observation variances are known, and $\sigma_u^2$ is the only hyperparameter left for estimation.

The posterior marginals are approximated directly. At each value of the hyperparameter, a Gaussian approximation to the field's conditional posterior is built at the mode, and a Laplace approximation to the hyperparameter's own posterior is formed from it. The field's marginals are then integrated over a design of hyperparameter points. The name refers to that nesting of one Laplace approximation inside another. We use the simplified Laplace strategy for the marginals and a central composite design for the integration.

We fit the differenced response $\tilde{\delta}_{d,g} = z_{d,g} - y_{p(d),g}$ as $\lambda_{p,g} + b_{p,g} + u_{d,g} + e_{d,g}$. The offsets $\lambda_{p,g}$ are province-by-stratum fixed effects under a vague Gaussian prior. The shared survey error $b_{p,g} \sim \mathcal{N}(0, \psi_{p,g})$ and the register error $e_{d,g} \sim \mathcal{N}(0, v_{d,g})$ both enter with their precisions held fixed at the known values, so $\lambda$'s posterior inherits $\psi_{p,g}$ at full strength and reproduces (20). The one estimated hyperparameter, the precision of $u_{d,g}$, is given a penalised-complexity prior (Simpson et al., 2017) under which $\Pr(\sigma_u > 1) = 0.01$. Lesotho is saturated at one area per province and stratum, so $\sigma_u^2$ is not identified there and its posterior returns its prior, which we report as such.

The stochastic partial differential equation representation of Lindgren, Rue and Lindström (2011) extends this machinery to continuously indexed Gaussian fields with Matérn covariance, by approximating them on a triangulated mesh by a sparse Gaussian Markov random field. Our units are areal and their precision is already sparse, so that construction is not needed here. It would be the route to an offset defined as a continuous surface, beyond the reach of these data.

##### A.4 Comparison of the three estimation routes

Table S1 gives the three routes at the resolution reported in the paper. The latent field is Gaussian, so the Laplace marginalisation is exact and the marginal-likelihood predictor is the correctly specified block-covariance EBLUP of (18), in which the linking errors share the province survey component $\psi_p$. The three differ materially only in $\sigma_u^2$. Below we explain first why they differ on it and then why that difference does not reach the printed results.

Table S1: The reported model (area × age × sex, block covariance) fitted three ways, by the measurement-error EBLUP of the main text, by the latent-field Laplace fit (TMB) and by a fully Bayesian fit (INLA). National selection odds ratios are inverse-variance-weighted over province–stratum cells throughout, so that the three routes are compared on one aggregation rule; they are therefore not the population-standardised figures reported in Section 5. Lesotho has one area per province and stratum, so $\sigma_u^2$ is not identified there.

| | units | | national selection OR | | | |
|---|---|---|---|---|---|---|
| Country | areas | cells | EBLUP | TMB | INLA | $\hat{\sigma}_u^2$ (TMB; EBLUP; INLA) |
| Mozambique | 159 | 56 | 1.544 | 1.544 | 1.544 | 0.216; 0.232; 0.234 |
| Zimbabwe | 63 | 57 | 2.855 | 2.848 | 2.856 | 0.137; 0.163; 0.169 |
| Malawi | 29 | 10 | 2.200 | 2.194 | 2.203 | 0.098; 0.112; 0.122 |
| Lesotho | 10 | 49 | 1.528 | 1.528 | 1.501 | not identified |

The shared survey error $\psi_p$ enters the two likelihood specifications differently. The Fay–Herriot differencing takes it into the block covariance (18), whereas the joint fit treats $\eta_p$ as a free fixed effect and conditions on it. That difference moves $\hat{\sigma}_u^2$ the same way in every country where the parameter is identified. TMB is below the EBLUP value and INLA above it in Mozambique, Zimbabwe and Malawi alike. The joint fit absorbs one free mean per province–stratum cell before $\sigma_u^2$ is estimated from the remaining variation, so it is subject to the incidental-parameter attenuation of roughly $(n_p - 1)/n_p$. With about 16, 6 and 6 areas per cell, that predicts shortfalls near 6, 16 and 17 per cent, against 7, 16 and 13 per cent observed. The direction is right in all three countries. The ordering between the two smaller countries is imperfect, as expected when $n_p$ varies within a country. The differenced fit never fully absorbs the province level, since $\log |V_p|$ retains $\log(1 + \psi_p a_p)$, a term that depends on $\sigma_u^2$ and rewards larger values, so it is the less attenuated of the two. INLA integrates over the province effects under priors and returns a posterior mean, which is the highest of the three. Zimbabwe's TMB value lies at the lower limit of the EBLUP profile interval, which a differently attenuated estimate from a different model need not respect.

A material disagreement on $\sigma_u^2$ nonetheless leaves the printed predictions unchanged. Refitting each country with the block-covariance EBLUP reproduces the latent-field district predictor. We make that comparison at the age-collapsed resolution of Appendix B, because those are the district predictions reported in this paper, whereas the age–sex fit of Section 5 supplies offsets and no printed district estimates. The comparison covers every unit of the three countries, with Harare and Bulawayo each pooled to one area in Zimbabwe (Appendix B). The maximum absolute difference on the logit scale is 0.009 in Mozambique, 0.004 in Zimbabwe and 0.003 in Malawi, with correlations above 0.9999. Lesotho enters at the admin-1 fit of Section 5, where one area per province leaves both routes returning the survey estimate, so its difference vanishes identically.

The reason is that $\sigma_u^2$ enters the predictor only through $\gamma_{d,g}$ and the weights forming $\hat{\lambda}_{p,g}$, and its effect on the weight is smallest where the weight is most extreme,

$$\frac{\partial \gamma_{d,g}}{\partial \sigma_u^2} = \frac{v_{d,g}}{(\sigma_u^2 + v_{d,g})^2},$$

which vanishes at both extremes of $v_{d,g}$ and peaks at $v_{d,g} = \sigma_u^2$. The two routes therefore agree most closely in the thickest cells and in the thinnest, where the weights approach one and zero together for any value of $\sigma_u^2$, and least closely in between. The reported fits are in the thick-cell regime, with median $\gamma_d$ from 0.991 to 0.996 over the whole range of $\sigma_u^2$ across the three routes in all three countries, so the disagreement on $\sigma_u^2$ does not reach the district predictions.

The INLA fit of Appendix A.3 retains the block covariance by construction, since the survey error enters as the cell-level term $b_{p,g}$ with its variance fixed at the known value, confounded with $\lambda_{p,g}$. Folding it into the observation variance instead would restore the diagonal working covariance and the understated intervals described below. The posterior standard deviation of $\lambda_{p,g}$ matches the Sherman–Morrison form (20) to within 1.3% in the worst cell of any country, against a 106 to 244% departure from the independent-error form. The national selection odds ratios agree to 0.6% or better in Mozambique, Zimbabwe and Malawi, and the INLA standard error is within about 1% of the EBLUP one. The posterior means of $\sigma_u^2$ are a little above the maximum-likelihood values, with credible intervals clear of zero, so the within-province variation of Section 5.1 does not depend on the estimation route. Lesotho is the instructive exception. It is saturated at one area per province, so its $\sigma_u^2$ posterior is its prior, and the fully Bayesian route integrates over that prior and returns a standard error 39% wider than the empirical-Bayes one. Where a variance component is not identified, the reported uncertainty depends on what is assumed about it, and we report the empirical-Bayes value with $\hat{\sigma}_u^2$ at zero.

Table S2 gives the latent-field offsets at the reported age–sex resolution, against the EBLUP values of Figure 1. No printed result depends on the choice among the three routes. We report the EBLUP route throughout and give all three here. Restricted maximum likelihood would be above the two likelihood values and is the natural response if the attenuation itself is at issue.

Table S2: Selection odds ratios by age and sex, latent-field (TMB) fit, the cross-check to Figure 1. Malawi is sex-only (women 1.81, men 2.77). Lesotho has one unit per province and stratum at this resolution, so the two routes coincide exactly.

| | 15–24 | | 25–49 | | 50+ | |
|---|---|---|---|---|---|---|
| Country | Female | Male | Female | Male | Female | Male |
| Mozambique | 1.25 | 1.61 | 1.48 | 1.76 | 1.96 | 1.72 |
| Zimbabwe | 2.86 | 4.31 | 2.08 | 3.89 | 2.44 | 3.55 |
| Lesotho | 1.27 | 2.28 | 1.29 | 2.45 | 0.84 | 2.21 |

An independent-error working covariance would leave the point estimates almost unchanged and the standard errors badly understated, for the reason set out at (20). Fitting the same data under an independent-error covariance narrows the standard error of the national selection odds ratio by a factor of 1.74, 1.60, 1.39 and 1.60 in Mozambique, Zimbabwe, Malawi and Lesotho, at the admin-2 fit of Appendix B. The factor is 1.00 at Lesotho's age–sex fit, where one area per province and stratum leaves the two working covariances identical. Under (20) instead, the differenced fit agrees with the latent-field fit, which conditions on the province level and so never lets the survey error average away. The two then agree on the national odds ratio to within about 1% and on its standard error to within 3% in every country, against the 1.39 to 1.74 fold disagreement under independent errors. What remains is the cost of holding $\hat{\sigma}_u^2$ fixed, which is propagated by the joint Hessian and omitted by the plug-in calculation. That part is the $O(1/m)$ Prasad–Rao correction (Prasad and Rao, 1990; Osgood-Zimmerman and Wakefield, 2023). Lesotho's age–sex fit has one unit per province and stratum, so nothing can average away and the two routes coincide there whichever covariance is used.

Table S3: Admin-2 (no age–sex) national selection odds ratios by fitting route, with $\hat{\sigma}_u$ the within-province SD on the logit scale (EBLUP fit). Every fit converges with positive-definite Hessian. Lesotho is fitted at its 72 community councils.

| Country | Areas | $\hat{\sigma}_u$ | OR (EBLUP) | OR (TMB) |
|---|---|---|---|---|
| Mozambique | 159 | 0.45 | 1.66 (1.45, 1.90) | 1.66 (1.45, 1.90) |
| Zimbabwe | 63 | 0.35 | 3.00 (2.55, 3.54) | 3.00 (2.56, 3.53) |
| Malawi | 29 | 0.33 | 2.12 (1.74, 2.57) | 2.11 (1.75, 2.56) |
| Lesotho | 72 | 0.33 | 1.22 (1.05, 1.41) | 1.22 (1.05, 1.41) |

##### A.5 A reference distribution for the transport test

The statistic $S^2_{p,g}$ of Section 4.3 is a weighted sum of squared normal variables, so it is right-skewed when few districts enter it. As a sensitivity check on the normal reference of (29), we match its first two moments to a scaled $\chi^2$, $S^2_{p,g} \approx c\,\chi^2_k$ with $k = 2\hat{\tau}^2_{p,g}/\hat{\mathrm{Var}}(S^2_{p,g})$ and $c = \hat{\mathrm{Var}}(S^2_{p,g})/(2\hat{\tau}_{p,g})$. Across the 23 tested provinces $k$ runs from 2.0 to 5.9, no province rejects transport at the one-sided 5% level, and the smallest $p$-value, for Mashonaland West, is 0.063 against 0.040 under the normal reference.

#### B Setting with minimal covariate structure

Where the routine records are available by district (admin-2) without age and sex, we calibrate at the district level with a province-level offset. Mozambique, Zimbabwe and Malawi are fitted at admin-2 (159, 63 and 29 districts, with Harare and Bulawayo each pooled to one area); Lesotho is fitted at its 72 community councils, one level below its LePHIA admin-1 design. The age-collapsed national selection odds ratios (Table S3) are close to the population-standardised age × sex × province figure of Section 5 in Mozambique (1.66 against 1.70) and below it in Malawi (2.12 against 2.36), Zimbabwe (3.00 against 3.67) and Lesotho (1.22 against 1.80). Collapsing the strata averages away the male cells with the strongest selection, and the shortfall is widest where the gap between men and women is widest (Table S2 in Appendix A), at 32% in Lesotho and 18% in Zimbabwe against 2% in Mozambique. The two fitting routes, the measurement-error EBLUP and the latent-field TMB, give national odds ratios within a few per cent of each other here, and their district predictors agree on the terms set out in Appendix A.

The calibrated district predictions, reported through the calibration identity as $\hat{\theta}_d = \mathrm{logit}^{-1}(z_d - \hat{\lambda}_{p(d)})$, span the expected range (Table S5), with median undiagnosed positivity from 1.0% in Malawi to 3.6% in Mozambique and district maxima up to 15.4%. The district-level variance decomposition attributes 82–99% of district prediction variance to the survey calibration (the $\hat{\lambda}$ term) and the small remainder to RHIS sampling noise. Under the transport assumption, the spatial resolution of the register is therefore obtained at little added uncertainty, and precision is limited by the survey at the calibration level. The share is lowest in Lesotho, whose community-council test volumes are smallest, so the register term is largest relative to the calibration. When $\sigma^2_\tau$ is set to its estimate $\hat{\sigma}^2_\tau$, the transport-failure term is the largest component of the survey-year variance at the median district of Table S6 (63% in Mozambique and 71% in Zimbabwe). Precision is then limited by the number of districts with survey estimates.

The same volumes set the shrinkage weight $\gamma_{d,g}$ of (26), and this fit is where it is closest to one. The parameter $\sigma^2_u$ is identified in all four countries here, so the shrinkage predictor and the calibrated register reported in Table S5 are nearly the same object (Table S4). Lesotho's weight is lowest for the same reason that its calibration share is lowest. Splitting the register by age and sex divides those counts and the weight falls with them, to 0.05 in Mozambique's thinnest cell and 0.46 in Zimbabwe's, so the weights span a factor of nineteen across Mozambique's cells.

The two predictors then separate by 0.16 in Mozambique and 0.10 in Zimbabwe at the ninetieth percentile. In Section 5, the age–sex fit is reported through its offsets and $\sigma_u^2$, so those separations affect no printed prediction.

Table S4: Median shrinkage weight $\gamma$ of (26) and the median absolute separation between the shrinkage predictor and the calibrated register, at the admin-2 fit of this appendix and at the age–sex fit of Section 5. Separations are on the logit scale, and are $(1-\gamma)$ times the gap between the survey estimate and the calibrated register, so they fall as $\gamma$ approaches one. Lesotho separates most at admin-2 because its community-council volumes are smallest and its $\gamma$ is lowest. Its age–sex fit has one unit per province and stratum, so no weight is defined there.

| | admin-2 | | age–sex | |
|---|---|---|---|---|
| Country | median $\gamma$ | separation | median $\gamma$ | separation |
| Mozambique | 0.995 | 0.002 | 0.95 | 0.012 |
| Zimbabwe | 0.992 | 0.001 | 0.94 | 0.012 |
| Malawi | 0.996 | 0.001 | 0.99 | 0.002 |
| Lesotho | 0.924 | 0.022 | saturated | |

Aggregated with population weights, the calibrated district estimates reproduce the survey's province estimates closely, though not exactly, because $\hat{\lambda}_p$ is a precision-weighted mean of district log-odds differenced against the province log-odds, and the log-odds of an average is not the average of log-odds. Weighting districts by census adult population, and Lesotho's community councils by WorldPop 2020 population, the mean gap between the aggregate and the survey estimate is +0.08, +0.05 and +0.02 percentage points in Mozambique, Zimbabwe and Malawi, with median absolute gaps of a quarter of the survey's standard error or less. Lesotho's councils aggregate below the survey in all ten districts, by 0.48 points on average, or 0.82 of the survey's standard error at the median, and a province-level benchmarking shift would remove that difference at negligible cost to the intervals.

Table S6 applies the interval (23) of Section 4.1 to the median district of each country with a transport diagnostic, for years after the survey. The transport term is the largest addition in the survey year itself, widening the Mozambique interval from 3.2 to 5.4 percentage points. The common drift then dominates the growth, and five years after the survey the interval is 8.4 points wide in Mozambique, 4.1 in Zimbabwe and 2.0 in Malawi, where $\hat{\sigma}_\tau^2$ is negative and set to zero.

Whether the area effect needs spatial structure is a separate question, and we examine it at this fit, whose units are the districts for which an adjacency graph is defined. Comparing independent, intrinsic conditional autoregressive (ICAR) and Besag–York–Mollié (BYM2) district fields by the deviance information criterion (DIC) gives no support for within-province spatial structure. The three specifications are separated by at most 0.8 DIC units in Mozambique and 0.1 in Malawi, and in Lesotho the ICAR field's DIC is 1.9 units higher than the independent field's, where lower values are preferred. Differences of this size are not evidence for the more elaborate field, so we keep the independent district effect $u_d$ of (16). Neighbours are resolved for 151 of Mozambique's 159 units, 26 of Malawi's 29 and all 72 of Lesotho's community councils. The remainder, the island districts of Ibo and Likoma, the seven urban districts of Maputo City and the two Mzimba divisions, enter as isolated nodes and receive no spatial smoothing, so the comparison is conservative toward the independent field. The comparison is made in Mozambique, Malawi and Lesotho only. Zimbabwe's routine extract reports Harare and Bulawayo on a health-administrative geography of central hospitals and city health units that occupy no contiguous area, and the adjacency graph has not yet been rebuilt for the pooled 63-district geography used here.

Table S5: District-level prediction summary, admin-2 fit, $\hat{\theta}_d = \text{logit}^{-1}(z_d - \hat{\lambda}_{p(d)})$ with $\text{Var}(\text{logit}\,\hat{\theta}_d - \text{logit}\,\theta_d) = v_d + \text{se}(\hat{\lambda}_{p(d)})^2$. "CI width" is the median 95% interval width under transport, and "with $\hat{\sigma}_\tau^2$" adds the transport-failure variance of Section 4.3 (Table 4; negative estimates set to zero, and none available for Lesotho); "% var calib." is the median share of the transport-only variance from $\hat{\lambda}$.

| Country | Areas | Median $\hat{\theta}$ | Range | CI width | with $\hat{\sigma}_\tau^2$ | % var calib. |
|---|---|---|---|---|---|---|
| Mozambique | 159 | 3.6% | 0.7–15.4% | 2.7 pp | 5.0 pp | 96% |
| Zimbabwe | 63 | 1.8% | 0.5–6.1% | 1.7 pp | 2.8 pp | 98% |
| Malawi | 29 | 1.0% | 0.3–2.4% | 0.8 pp | 0.8 pp | 99% |
| Lesotho | 72 | 2.3% | 0.9–6.0% | 1.9 pp | – | 82% |

Table S6: Width of the 95% interval (23) for the calibrated register estimate at each country's median district, $h$ years after the last survey, in percentage points. Columns add terms cumulatively: the survey-year variance $v + \text{Var}(\hat{\lambda})$ of (22), the transport term $\hat{\sigma}_\tau^2$, the diffusion $h\hat{\sigma}_\Delta^2$ and the squared common drift $h^2\hat{\mu}^2$, with $\hat{\sigma}_\Delta^2 = 0.0045$ and $|\hat{\mu}| = 0.071$ per year from the pooled two-round data. The register term is held at its survey-year value.

| Country | $\hat{\theta}$ (%) | $h$ | $v + \text{Var}(\hat{\lambda})$ | $+\hat{\sigma}_\tau^2$ | $+h\hat{\sigma}_\Delta^2$ | $+h^2\hat{\mu}^2$ |
|---|---|---|---|---|---|---|
| Mozambique | 3.6 | 0 | 3.2 | 5.4 | 5.4 | 5.4 |
| | | 1 | 3.2 | 5.4 | 5.5 | 5.7 |
| | | 3 | 3.2 | 5.4 | 5.7 | 6.7 |
| | | 5 | 3.2 | 5.4 | 5.9 | 8.4 |
| Zimbabwe | 1.8 | 0 | 1.3 | 2.5 | 2.5 | 2.5 |
| | | 1 | 1.3 | 2.5 | 2.5 | 2.6 |
| | | 3 | 1.3 | 2.5 | 2.6 | 3.2 |
| | | 5 | 1.3 | 2.5 | 2.7 | 4.1 |
| Malawi | 1.0 | 0 | 1.0 | 1.0 | 1.0 | 1.0 |
| | | 1 | 1.0 | 1.0 | 1.0 | 1.1 |
| | | 3 | 1.0 | 1.0 | 1.1 | 1.4 |
| | | 5 | 1.0 | 1.0 | 1.2 | 2.0 |

### C The generative model

Figure S1 shows the epidemiology produced by the generative model of Section 6.1, and Table S7 gives every parameter of the model.

Positivity among people not yet diagnosed depends on prevalence and on awareness of status together. The two vary across strata in the same direction, with prevalence highest in older women and awareness also higher in women and at older ages, and people who become aware of their status leave the group not yet diagnosed. The undiagnosed burden therefore shifts toward men and the young even where prevalence peaks in older women. The generative model reproduces both quantities, so that the age–sex structure of selection found in Section 5 is tested against a population that resembles the real one.

The unscaled district standard deviations give $\sigma_u^2 = 0.05$. Because the scaling factor is common, the share of $\sigma_u^2$ due to transport failure of the offset stays fixed at 34%. The diagnostic of Section 4.3 puts that share at 44% in Mozambique and 64% in Zimbabwe and below zero in Malawi, each with an interval spanning zero and the whole of $\sigma_u^2$, and Section 6.2 reports the results across shares from 20 to 80%.

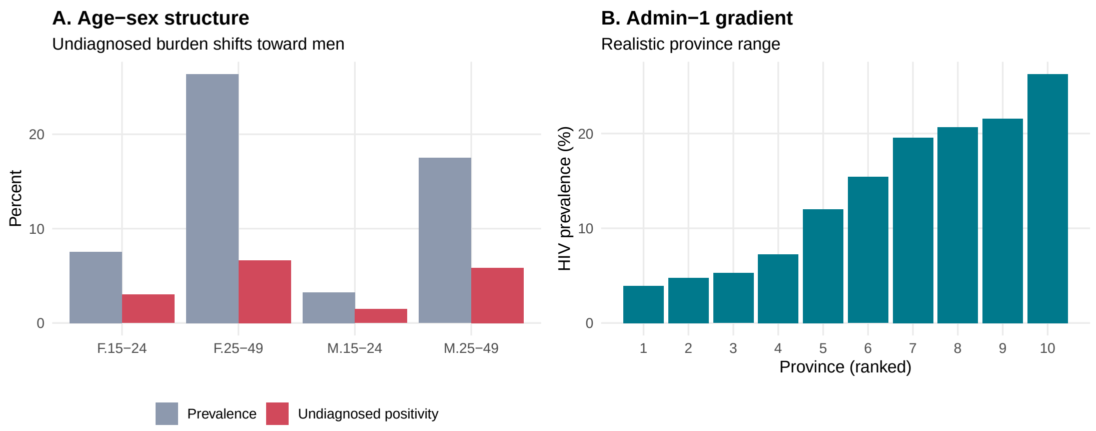


Figure S1: Survey-realistic epidemiology in the generative model. **A**: the undiagnosed burden shifts toward men even though prevalence peaks in older women. **B**: province (admin-1) prevalence spans a realistic gradient.

#### Offset scenarios

The offset baseline is the stratum value of Table S7 plus a district term, and four scenarios add time dynamics to it. Under the constant scenario the offset does not move, so the calibration derived from one survey remains valid throughout the period. Under drift, the offset of each unit changes by a constant amount at every wave, drawn separately for each unit with standard deviation 0.10 per wave. Under shock, a common break of 0.50 on the log-odds scale arrives at wave 3 and persists, as a stylised interruption to testing funding or the arrival of a new testing modality. Under reshuffle, the offset follows a stationary first-order autoregression in each unit, with $\phi = 0.30$ and innovation standard deviation 0.25, so areas change their relative order between waves. The coverage figures quoted in Section 6.2 are given under each of these four, and Figure S2 shows the offset and the estimand under each. The estimand's path is the same under all four.

The temporal dynamics of the offset are set as scenarios and are not calibrated to the two-round movement of Section 5.2, since that movement is measured on the PEPFAR indicators, a different source from the ministry register. The cross-sectional sizes in Table S7 come from the four-country fit of Section 5.

Table S7: Parameters of the generative model of Section 6.1. Stratum entries are ordered women 15–24, women 25–49, men 15–24, men 25–49. Province effects are an ordered gradient across the ten provinces scaled to a standard deviation of 0.85, with independent jitter of standard deviation 0.10 added.

| Symbol | Meaning | Value |
|---|---|---|
| $P$ | provinces (admin-1) | 10 |
| | districts per province | 4 |
| $D$ | districts | 40 |
| $T$ | annual waves | 6 |
| | age–sex strata | 4 |
| | population share by stratum | 0.27, 0.26, 0.26, 0.21 |
| $\bar{q}_g$ | baseline prevalence by stratum | 0.060, 0.240, 0.025, 0.150 |
| $\bar{a}_g$ | baseline awareness by stratum | 0.62, 0.82, 0.55, 0.72 |
| $\rho$ | prevalence decline per wave (probability scale) | 0.035 |
| $\kappa$ | awareness gain per wave (logit scale) | 0.12 |
| $\beta_p$ | province gradient on logit prevalence | 0.85 |
| $\gamma_d$ | district effect on logit prevalence (sd) | 0.37 |
| $\gamma'_d$ | district effect on logit awareness (sd) | 0.31 |
| $\lambda^0$ | offset baseline by stratum (log-odds) | 0.55, 0.35, 1.00, 0.80 |
| | district effect on the offset (sd) | 0.31 |
| $\sigma^2_u$ | within-province variance of the district residual | 0.20 |
| $v$ | register noise variance on the logit | 0.010 |
| | survey design effect | 2.0 |
| $n$ | sampled per stratum per surveyed district-wave | 120 |

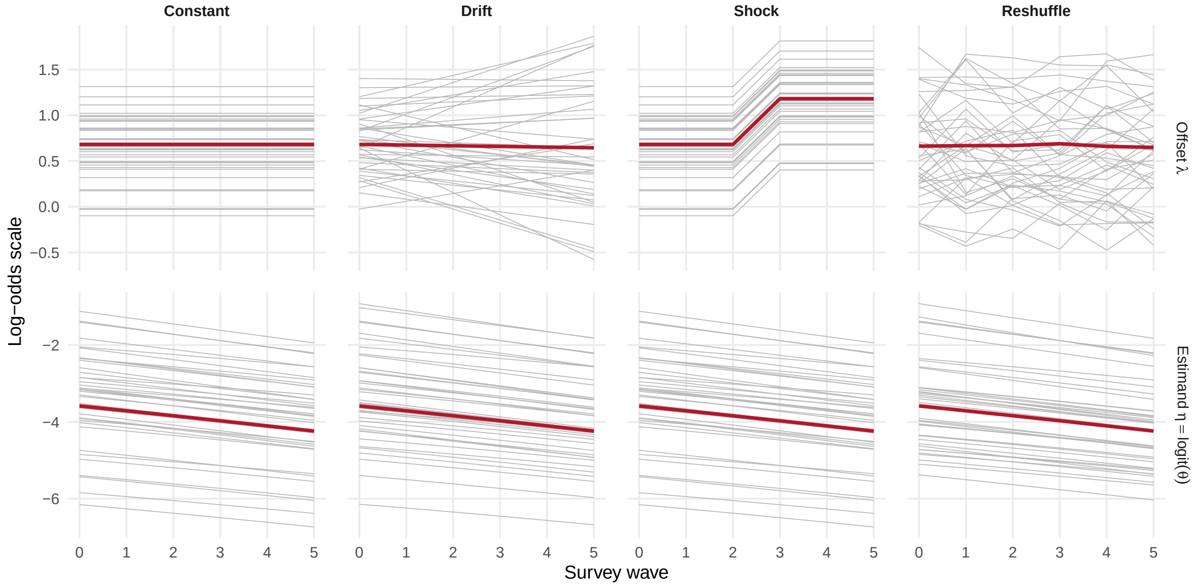


Figure S2: The offset $\lambda$ and the estimand $\eta = \operatorname{logit}\theta$ under all four offset scenarios, from one simulated country at a common seed. Grey lines are 40 of the 160 district by age–sex units, and the red line is the mean over all of them. Only the offset is perturbed under the scenarios. In the generative model $\eta$ is built from prevalence and awareness before the offset dynamics are applied, so the lower row is the same path four times.

Table S8: Accuracy against the within-province variance $\sigma_u^2$ under each offset scenario, with the audit of Table 7 and one panel per reporting level. Entries are root-mean-squared errors in percentage points, averaged over 150 replications, for the register alone (Reg.), the survey audit alone (Surv.) and the calibrated estimate (Cal.). $\sigma_u^2$ is varied by scaling the district standard deviations of prevalence, awareness and the offset by a common factor, so the share of $\sigma_u^2$ due to transport failure of the offset stays fixed. The age-collapsed estimates for the four countries are 0.108, 0.112, 0.126 and 0.202, the main-fit values 0.11 to 0.23, and Table 7 uses 0.20. In bold, the lowest error in each scenario and row.

| | Constant | | | Drift | | | Shock | | | Reshuffle | | |
|---|---|---|---|---|---|---|---|---|---|---|---|---|
| $\sigma_u^2$ | Reg. | Surv. | Cal. | Reg. | Surv. | Cal. | Reg. | Surv. | Cal. | Reg. | Surv. | Cal. |
| National | | | | | | | | | | | | |
| 0.05 | 2.59 | 0.84 | **0.44** | 2.74 | 0.84 | **0.53** | 4.00 | 0.84 | **0.44** | 2.74 | 0.84 | **0.52** |
| 0.10 | 2.67 | 0.87 | **0.49** | 2.82 | 0.87 | **0.58** | 4.10 | 0.87 | **0.49** | 2.83 | 0.87 | **0.57** |
| 0.20 | 2.84 | 0.95 | **0.60** | 2.99 | 0.95 | **0.69** | 4.31 | 0.95 | **0.60** | 2.99 | 0.95 | **0.68** |
| 0.30 | 3.00 | 1.02 | **0.70** | 3.15 | 1.02 | **0.79** | 4.51 | 1.02 | **0.70** | 3.15 | 1.02 | **0.78** |
| 0.50 | 3.31 | 1.17 | **0.90** | 3.46 | 1.17 | **1.00** | 4.89 | 1.17 | **0.90** | 3.46 | 1.17 | **0.99** |
| Admin-1 | | | | | | | | | | | | |
| 0.05 | 3.14 | 2.28 | **0.61** | 3.33 | 2.28 | **0.75** | 4.81 | 2.28 | **0.61** | 3.34 | 2.28 | **0.74** |
| 0.10 | 3.28 | 2.34 | **0.73** | 3.47 | 2.34 | **0.86** | 4.96 | 2.34 | **0.73** | 3.47 | 2.34 | **0.86** |
| 0.20 | 3.54 | 2.46 | **0.95** | 3.73 | 2.46 | **1.07** | 5.27 | 2.46 | **0.95** | 3.73 | 2.46 | **1.07** |
| 0.30 | 3.80 | 2.59 | **1.16** | 3.98 | 2.59 | **1.28** | 5.56 | 2.59 | **1.16** | 3.99 | 2.59 | **1.28** |
| 0.50 | 4.29 | 2.85 | **1.56** | 4.47 | 2.85 | **1.68** | 6.11 | 2.85 | **1.56** | 4.47 | 2.85 | **1.68** |
| District | | | | | | | | | | | | |
| 0.05 | 3.30 | 2.66 | **0.84** | 3.57 | 2.66 | **1.08** | 4.97 | 2.66 | **0.84** | 3.57 | 2.66 | **1.08** |
| 0.10 | 3.57 | 2.80 | **1.08** | 3.84 | 2.80 | **1.30** | 5.27 | 2.80 | **1.08** | 3.84 | 2.80 | **1.30** |
| 0.20 | 4.10 | 3.09 | **1.52** | 4.36 | 3.09 | **1.71** | 5.86 | 3.09 | **1.52** | 4.36 | 3.09 | **1.72** |
| 0.30 | 4.60 | 3.37 | **1.92** | 4.86 | 3.37 | **2.11** | 6.42 | 3.37 | **1.92** | 4.86 | 3.37 | **2.11** |
| 0.50 | 5.56 | 3.92 | **2.66** | 5.80 | 3.92 | **2.85** | 7.47 | 3.92 | **2.66** | 5.80 | 3.92 | **2.84** |
| District × age–sex | | | | | | | | | | | | |
| 0.05 | 3.84 | 3.42 | **1.35** | 4.39 | 3.42 | **1.86** | 5.61 | 3.42 | **1.35** | 4.41 | 3.42 | **1.89** |
| 0.10 | 4.13 | 3.57 | **1.58** | 4.67 | 3.57 | **2.07** | 5.92 | 3.57 | **1.58** | 4.68 | 3.57 | **2.09** |
| 0.20 | 4.66 | 3.89 | **2.00** | 5.20 | 3.89 | **2.48** | 6.52 | 3.89 | **2.00** | 5.21 | 3.89 | **2.49** |
| 0.30 | 5.18 | 4.20 | **2.41** | 5.71 | 4.20 | **2.87** | 7.09 | 4.20 | **2.41** | 5.72 | 4.20 | **2.89** |
| 0.50 | 6.16 | 4.81 | **3.19** | 6.68 | 4.81 | **3.64** | 8.15 | 4.81 | **3.19** | 6.67 | 4.81 | **3.65** |

Table S9: Margin of the calibrated estimate over the survey audit, in percentage points of root-mean-squared error, against the share of the within-province variance $\sigma_u^2 = 0.20$ due to transport failure of the offset, under the constant and drift scenarios, with the audit of Table 7. The share is varied by scaling the district standard deviation of the offset against those of prevalence and awareness, and 0.34 is the share used elsewhere. A negative margin means the survey alone is more accurate.

| | Constant | | | | Drift | | | |
|---|---|---|---|---|---|---|---|---|
| Share | National | Admin-1 | District | Cell | National | Admin-1 | District | Cell |
| 0.20 | 0.46 | 1.73 | 2.01 | 2.31 | 0.37 | 1.60 | 1.78 | 1.79 |
| 0.34 | 0.36 | 1.53 | 1.60 | 1.92 | 0.27 | 1.40 | 1.40 | 1.44 |
| 0.44 | 0.29 | 1.40 | 1.34 | 1.65 | 0.20 | 1.28 | 1.15 | 1.20 |
| 0.64 | 0.15 | 1.15 | 0.86 | 1.16 | 0.06 | 1.03 | 0.68 | 0.74 |
| 0.80 | 0.05 | 0.97 | 0.51 | 0.79 | −0.04 | 0.85 | 0.34 | 0.39 |

##### Audit and replication

At each wave, $k$ of the $D = 40$ districts are drawn for the audit uniformly at random without replacement, independently across waves, and a surveyed district contributes all four of its strata. In Section 6.2, $k = 8$, and the results are averages over 150 replications, as are the coverage figures, at seeds $5000 + r$ and $6000 + r$ for replication $r$. In the sweeps behind Sections 6.3 and 6.4, we use the same audit of 8 districts per wave, since only the offset varies in them. An audit of a given size can be placed by other rules, including one that revisits the districts whose offset is least well determined, and those rules belong with the design question of Section 7.3.

##### The estimators

The simulated offset has no province component, so the broad area of (11) is the stratum nationally. The estimate $\hat{\lambda}_{gt}$ is the generalised least squares mean of $z - \hat{Z}$ over the $m = 8$ audited districts of stratum $g$ at wave $t$, weighted by $(\hat{\sigma}_\tau^2 + \psi_{it} + v)^{-1}$. Each audited district has its own survey error, so no survey term is shared, and $\hat{\sigma}_\tau^2$ is estimated by restricted maximum likelihood over the wave's audited cells. Intervals use $v + \mathrm{Var}(\hat{\lambda}_{gt}) + \hat{\sigma}_\tau^2$, the survey-year case of (23).

##### Coverage of the calibrated intervals

The 95% intervals of Section 6.2 use $v + \mathrm{Var}(\hat{\lambda}_{gt}) + \hat{\sigma}_\tau^2$. Their coverage is similar in audited cells (0.919 to 0.927) and elsewhere (0.890 to 0.903). With $\sigma_\tau^2$ set to its generating value, coverage is 0.955 under the constant and shock offsets. Under drift and reshuffle the offset's spread widens over the waves beyond that value and coverage stays near 0.90 either way. Without the $\sigma_\tau^2$ term, coverage falls to between 0.795 and 0.848.

##### Within-province variance and transport failure

The within-province variance $\sigma_u^2$ is the subject of the transport assumption, and the results of Section 6.2 are read at 0.20, between the main-fit values of Zimbabwe and Mozambique. When it is varied from 0.05 to 0.50, a range whose upper end exceeds every value observed in these data, the calibrated estimate remains more accurate than the audit at every reporting level (Table S8). The margin narrows as areas within a province differ more, from 1.66 to 1.29 percentage points at admin-1 under a constant offset, and it is smallest at national level throughout. Holding $\sigma_u^2$ at 0.20 and moving the share due to transport failure from 20 to 80% narrows the district margin from 2.01 to 0.51 points under a constant offset and from 1.78 to 0.34 under drift (Table S9). At 64%, Zimbabwe's point estimate, the margin is 0.86 and 0.68. The calibrated estimate stays ahead of the audit at every subnational level across that range, because the survey-only estimate in an unaudited cell is the stratum mean over the audited districts, a cruder benchmark than the province mean assumed in condition (30).

##### The drift sweep

The annual drift of the offset is expressed as a fraction of its spread across areas within one year, and varied from 0 to 1. A quantity that varies by a given amount across areas is unlikely to vary by more than that in one place within one year. This spread can also be estimated from a single survey round, whereas separating a sustained drift from a one-off shift requires at least three rounds.

##### A register without age and sex detail

Table S10 gives the cost of pooling at each country's spread. Malawi's figure is not comparable with the others, because its register records sex without age, so its spread measures the male–

Table S10: The cost of aggregating over age and sex, with each country evaluated at its own between-stratum spread of the offset. Entries are the root-mean-squared error of estimated undiagnosed positivity in percentage points, taken across all cells of one simulated population and averaged over 150 replications. In this sweep we vary the between-stratum spread directly and hold the offset otherwise fixed, so the four offset scenarios of Appendix C have no role in this sweep. A positive gain means pooling the strata is the more accurate choice at that spread, and the two are equal at a spread of 0.30. Malawi's spread measures the male–female contrast alone, because its register records sex without age.

| Country | Between-stratum spread | Stratum-resolved | One offset for all strata | Gain from pooling |
|---|---|---|---|---|
| Mozambique | 0.16 | 2.00 | 1.77 | +0.23 |
| Zimbabwe | 0.28 | 2.00 | 1.97 | +0.04 |
| Malawi | 0.30 | 2.00 | 2.01 | −0.00 |
| Lesotho | 0.43 | 2.00 | 2.32 | −0.32 |

female contrast alone. Whether the pooled calibration still improves on its two sources is a separate question, and this setting is its hardest test. We score the pooled estimate against the uncalibrated register and against the province survey mean on the same populations. Its root-mean-squared error is 2.45 to 3.25 percentage points lower than that of the register alone and 1.08 to 2.20 lower than that of the survey alone, at every spread on the grid, and the gain over the survey narrows as selection differs more between strata. The spread used here is the standard deviation of the offset across the strata of one area, a different quantity from the marginal age–sex contrasts of Section 5.1, which are averages over provinces.

##### The simulated truth

The simulated population is within the range present across the four countries, so the errors reported in Section 6 can be read against the application. National undiagnosed positivity falls from 4.58% at wave 0 to 2.40% at wave 5 and averages 3.41%, against 1.1% to 4.0% across the four surveys of Section 2.2. Adult prevalence falls from 13.9% to 11.7% and awareness of status rises from 67.2% to 78.3%. The implied selection odds ratio averages 2.15 at wave 0, with cells running from 0.70 to 5.82, against national figures of 1.70 to 3.67 in Section 5.1. Root-mean-squared errors given in percentage points in Section 6 should be read against these levels, which differ by resolution. The national figure is about 3%, while individual district by age–sex cells run from 0.11% to 25.99% across the waves, so the errors reported at the finest resolution are compared with a far wider spread of true values than the national ones. These figures are averaged over the same 150 replications as in Section 6.2. The four scenarios perturb the offset alone, so the simulated epidemiology is the same under each of them.
